# Complementary feeding practices in 80 low- and middle-income countries: prevalence and socioeconomic inequalities in dietary diversity, meal frequency and dietary adequacy

**DOI:** 10.1101/2020.12.01.20241372

**Authors:** Giovanna Gatica-Domínguez, Paulo A. R. Neves, Aluísio J. D. Barros, Cesar G. Victora

**Affiliations:** International Center for Equity in Health, Post-graduate Program in Epidemiology, Federal University of Pelotas, Brazil

## Abstract

**Objective:** To describe patterns and socioeconomic inequalities in complementary feeding practices among children aged 6-23 months in 80 low and middle-income countries (LMICs).

**Methods:** We analyzed national surveys carried out since 2010. Complementary feeding indicators for children aged 6-23 months included minimum dietary diversity (MDD), minimum meal frequency (MMF) and minimum acceptable diet (MAD). Between- and within-country inequalities were documented using relative (wealth deciles) and absolute (estimated household income) socioeconomic indicators.

**Results:** Only 21.3%, 56.2% and 10.1% of the 80 countries showed prevalence levels above 50% for MDD, MMF and MAD, respectively. Western & Central Africa showed the lowest prevalence for all indicators, whereas the highest for MDD and MAD was Latin America & Caribbean, and for MMF in East Asia & the Pacific. Log per capita gross domestic product was positively associated with MDD (R2 = 48.5%), MMF (28.2%) and MAD (41.4%). Pro-rich within-country inequalities were observed in most countries for the three indicators; pro-poor inequalities were observed in two countries for MMF, and in none for the other two indicators. Breastmilk was the only type of food with a pro-poor distribution, whereas animal-source foods (dairy products, flesh foods and eggs) showed the most pronounced pro-rich inequality. Dietary diversity improved sharply when absolute annual household incomes exceeded about US$20,000. There were no consistent differences among boys and girls for any of the indicators studied.

**Conclusion:** Monitoring complementary feeding indicators in the world and implementing policies and programs to reduce wealth-related inequalities are essential to achieve optimal child nutrition.

## INTRODUCTION

During the first two years of life, all children must be optimally breastfed and receive an appropriate and diverse diet from six months of age in order to achieve optimal growth and development^1–4^. Departures from optimal growth vary in different groups of countries. In most high-income countries (HIC), childhood overweight/obesity is a major concern, partially caused by high-energy-dense diets^5^. In low- and middle-income countries (LMIC), stunting (low height-for-age) and micronutrient deficiencies are more prevalent due to poor-quality diets^6^.

The introduction of a healthy diet in early childhood contributes to better food preferences and health outcomes throughout the life course^6^. In 2007, the World Health Organization (WHO) proposed a set of complementary feeding indicators for monitoring infant and young child feeding (IYCF) practices among children aged 6-23 months^7,8^. The core indicators address the diversity (minimum dietary diversity or MDD) and frequency (minimum meal frequency or MMF) of child diets. A third indicator - minimum acceptable diet or MAD – relates to child diets that met both diversity and frequency requirements. Analyses conducted in South Asia using national surveys found that children whose diets complied with the IYCF recommendations were less likely to be ill or malnourished^9,10^.

Socioeconomic inequalities represent a major threat to optimal feeding practices.^11,12^ Using the 2007 definitions, a UNICEF report analyzed data from up to 87 national surveys in LMICs. The study found low overall prevalence levels: 29.4% for MDD, 52.2% for MMF, and only 16% for MAD. The report also found that prevalence of the three indicators increased with household wealth within countries^13^.

In 2018, UNICEF and WHO updated the definitions of the three indicators, mainly in order to refine analyses of the diets of breastfed children^14^. A recent publication based on 49 national surveys from LMICs reported on MDD prevalence using the 2018 definition, but relied on the 2007 definition for measuring MMF, and MAD was calculated as the combination of these two. The regions with the lowest and highest proportions of children aged 6-23 months that met the three complementary feeding indicators requirements were Sub-Saharan Africa and Latin America & Caribbean, respectively. MDD prevalence ranged from 18% to 54%, MMF from 41% to 72%, and MAD from 9% to 40%. Stark disparities by wealth quintile were observed in most LMICs studied, particularly for MDD, which was also positively associated with GNI PPP at country level^15^. As far as we are aware, there are no multi-country analyses using the 2018 definitions of the three indicators.

The Sustainable Development Goals, part of the 2030 Agenda for Sustainable Development^16^ call for action towards a better future for all, which includes appropriate diets for children, addressing goals 2 (zero hunger) and 3 (good health and well-being). Disaggregated analysis by socioeconomic indicators, including recent nationally representative surveys carried out in LMICs, are essential to track progress and identify challenges regarding complementary feeding practices. In the present analyses, we describe wealth-related inequalities in complementary feeding practices among children aged 6-23 months in 80 LMICs. We used the 2018 definitions and provide breakdowns by wealth deciles, to allow greater granularity than wealth quintiles, and also by estimated absolute income of households in international dollars.

## METHODS

The database of the International Center for Equity in Health (www.equidade.org) includes over 400 national surveys with information on child health and nutrition in LMICs. We selected the most recent survey in each country, carried out since 2010, that included information on the three complementary feeding indicators described below, and sample sizes of at least 25 children aged 6-23 months in each wealth decile. A total of 80 surveys were included, being 41 Demographic Health Surveys (DHS; https://dhsprogram.com), 38 Multiple Indicator Cluster Surveys (MICS; http://mics.unicef.org/) and one modified version of the DHS from Ecuador (Encuesta Nacional de Salud y Nutrición 2012). DHS and MICS are highly comparable in terms of methodology and measurement protocols, allowing for the comparability of results^17^. All surveys rely on multistage sampling procedures, selecting regions within countries, administrative units within each region (e.g., municipalities), census tracts within each administrative unit, and households within each tract. All women aged 15-49 years from selected households are invited for an interview on the nutrition and health of their under-five children. Further information on survey methodology is available in each survey’s published national reports.

### Complementary feeding indicators

Three complementary feeding indicators were estimated for children aged 6-23 months, based on 24-hour dietary recall.^14^ MDD was calculated at the proportion of children who consumed foods and beverages from five or more out of eight food groups (see below). MMF was calculated as the number of breastfed children who consumed solid, semi-solid or soft foods the at least twice (if aged 6 - 8 months) or three times (if aged 9 - 23 months), plus the number of non-breastfed children who received at least four feeds during the previous day (including at least one feed of solid, semi-solid or soft foods); the resulting sum is divided by the number of children aged 6-23 months. Lastly, MAD was calculated as the percentage of children with satisfactory MDD and MMF, and are either breastfed or had at least two non-human milk feeds in the previous 24 hours.

For each of the eight food groups used to calculate the MDD indicator, we also reported the percentage of children who during the previous day consumed foods or beverages from each of the eight food groups: 1) breastmilk; 2) cereals and grains (grains, white/pale starchy roots, tubers, and plantains); 3) legumes and nuts (beans, peas, lentils, nuts, and seeds); 4) dairy products (milk, infant formula, yogurt, cheese); 5) flesh foods (meat, fish, poultry, organ meats); 6) eggs; 7) vitamin-A rich fruits and vegetables; and, 8) other fruits and vegetables. Although two surveys (Papua New Guinea 2016 and Guyana 2014) did not collect data on yogurt, we decided to proceed with calculation of the MMF indicator because of the reportedly low frequency of consumption compared to other dairy products.

### Socioeconomic indicators and analyses

#### Wealth deciles

Household asset scores, generated through principal component analysis (PCA), were available in the DHS and MICS datasets. The PCA includes variables on household assets, building materials, and utilities like water and electricity, which are adjusted for the place of residence.^18^ The first component of the PCA, a continuous variable, was used to classify households into wealth deciles, with the first decile (D1) representing the poorest 10% of all families and the tenth decile (D10) representing the wealthiest 10% of all families.

#### Per capita gross domestic product (GDP)

This indicator is expressed in current international dollars converted by purchasing power parity (PPP), is the sum of the gross value added by all resident producers in the country plus any product taxes and minus any subsidies not included in the value of the products. The PPP conversion factor is a spatial price deflator and currency converter that eliminates the effects of the differences in price levels between countries.^19^ We obtained the GDP data through the wbopendata module 16.3, which draws from the core World Bank development indicators. It presents the most current and accurate global development data available, compiled from officially-recognized international sources^20^.

#### Absolute income for each wealth decile

This was calculated based on the national income levels obtained from the World Bank database (http://databank.worldbank.org/data/reports.aspx?source=world-developmentindicators), and national income inequality data collected from the Standardized World Income Inequality Database (https://www.wider.unu.edu/project/wiidworld-income-inequality-database). Dollar values (2011 purchase power parity adjusted international dollars) were then assigned to each household wealth decile, accounting for income’s log-normal distribution^21^.

#### Slope index of inequality (SII)

The SII is a summary measure of absolute inequality, which is calculated through logistic regression models with the natural logarithms of the odds of the complementary feeding variables as the outcomes, and the wealth deciles as the independent variable. The SII represents the difference in the fitted value of the outcome between the highest and the lowest values of the wealth index scale^22^, and is interpreted as percentage points (p.p.).

### Statistical analysis

For each country included in the analyses, we estimated the prevalence of the three indicators at national level, by sex of the child and by wealth decile, and calculated the SII and its 95% confidence interval (CI). Next, we grouped the countries according to UNICEF world regions and World Bank income group classifications for the year of the survey^23,24^. Regional and income group estimates were weighted by the size of the population of children aged 6 to 23 months in the year when the survey was conducted^25^. Equiplot graphs were used to illustrate how weighted mean prevalence varied by wealth deciles.

We fitted polynomial equations for each complementary feeding indicator according to log GDP at national level, but this procedure did not improve the fit of the model compared to a linear equation. We present R^2^ values for the linear models, which express the proportion of the variance of the complementary feeding indicators that are explained by GDP. Scatter plots were used to graph the associations. Fractional polynomials were used to describe non-linear associations in graphical form of complementary feeding practices and absolute income. We also plotted within-country inequalities for the top three and bottom three countries in terms of each complementary feeding prevalence. Three countries (i.e., Papua New Guinea 2016, Sao Tome and Principe 2014, and Yemen 2013) had no available information on absolute income; hence they were excluded only for the absolute income analysis. Lastly, the prevalence of consumption of each of the eight food groups was calculated for the poorest and wealthiest deciles in each country and then grouped by world region.

All the analyses were performed in Stata 16.0 (StataCorp, College Station, TX, USA) considering the survey design, sampling weights, clustering, and stratification, and R version 3.6.0 for the absolute income graphs. The data used in our analyses are publicly available, and the institutions that conducted the surveys in each country handled the respective ethical clearance.

## RESULTS

The most recent surveys were analyzed for 80 countries, with dates ranging from 2010 to 2019 (median 2016). The number of children aged 6-23 months in the national surveys ranged from 332 in Montenegro to 71,762 in India, with a median of 2,581 children. Supplementary table 1 shows the surveys included in the analyses and sample sizes.

**Table 1.** Weighted mean prevalence of complementary feeding indicators for all children aged 6-23 months and stratified by sex, according to the regions of the world

| World region | All |  | Boys |  | Girls |  |
| --- | --- | --- | --- | --- | --- | --- |
|  | Mean | 95% CI | Mean | 95% CI | Mean | 95% CI |
| Minimum dietary diversity |  |  |  |  |  |  |
| Western & Central Africa | 19.0 | 16.6; 21.5 | 19.1 | 16.7; 21.6 | 19.0 | 16.5; 21.4 |
| Eastern & Southern Africa | 24.0 | 19.9; 28.1 | 24.1 | 20.1; 28.0 | 24.1 | 19.8; 28.3 |
| Middle East & North Africa | 36.2 | 26.5; 45.9 | 36.2 | 25.8; 46.7 | 36.3 | 27.4; 45.2 |
| Eastern Europe & Central Asia | 50.3 | 36.5; 64.0 | 50.2 | 36.3; 64.2 | 50.5 | 36.6; 64.4 |
| South Asia | 20.8 | 13.8; 27.7 | 20.5 | 13.1; 27.9 | 21.1 | 14.6; 27.5 |
| East Asia & the Pacific | 50.6 | 42.2; 59.0 | 49.9 | 42.2; 57.6 | 51.4 | 42.1; 60.6 |
| Latin America & Caribbean | 56.5 | 48.9; 64.1 | 53.5 | 46.1; 61.0 | 59.3 | 51.2; 67.4 |
| *All regions* | 27.1 | 23.9; 30.3 | 26.8 | 23.7; 29.9 | 27.5 | 24.2; 30.8 |
| Minimum meal frequency |  |  |  |  |  |  |
| Western & Central Africa | 39.6 | 36.4; 42.7 | 39.0 | 35.8; 42.2 | 40.1 | 37.0; 43.3 |
| Eastern & Southern Africa | 45.1 | 40.0; 50.3 | 44.9 | 39.8; 50.0 | 45.3 | 40.1; 50.5 |
| Middle East & North Africa | 62.9 | 56.6; 69.3 | 63.3 | 56.8; 69.8 | 62.5 | 56.2; 68.8 |
| Eastern Europe & Central Asia | 68.8 | 56.2; 81.5 | 68.1 | 55.7; 80.5 | 68.7 | 55.8; 81.7 |
| South Asia | 43.2 | 28.1; 58.3 | 43.7 | 28.5; 58.9 | 42.6 | 27.6; 57.6 |
| East Asia & the Pacific | 72.5 | 65.5; 79.5 | 73.1 | 66.2; 80.1 | 71.8 | 64.7; 79.0 |
| Latin America & Caribbean | 71.5 | 63.9; 79.0 | 71.7 | 64.3; 79.2 | 71.2 | 63.5; 78.9 |
| *All regions* | 48.7 | 45.2; 52.2 | 48.8 | 45.3; 52.3 | 48.5 | 45.1; 51.9 |
| Minimum acceptable diet |  |  |  |  |  |  |
| Western & Central Africa | 9.4 | 8.3; 10.6 | 9.0 | 7.9; 10.1 | 9.9 | 8.6; 11.1 |
| Eastern & Southern Africa | 13.3 | 10.6; 16 | 13.5 | 10.6; 16.3 | 13.1 | 10.5; 15.7 |
| Middle East & North Africa | 25.4 | 17.4; 33.5 | 25.6 | 16.9; 34.3 | 25.2 | 17.9; 32.6 |
| Eastern Europe & Central Asia | 37.7 | 23.2; 52.2 | 37.2 | 22.7; 51.6 | 38.2 | 23.2; 53.3 |
| South Asia | 12.1 | 5.1; 19.2 | 12.2 | 5.0; 19.4 | 12.1 | 5.1; 19.1 |
| East Asia & the Pacific | 39.7 | 31.5; 48.0 | 39.6 | 31.7; 47.5 | 39.9 | 31.3; 48.5 |
| Latin America & Caribbean | 43.7 | 36.4; 50.9 | 41.4 | 34.2; 48.5 | 45.7 | 38.0; 53.4 |
| *All regions* | 17.3 | 14.4; 20.2 | 17.1 | 14.3; 19.9 | 17.5 | 14.6; 20.4 |

**NOTE:** All estimates were weighted by the size of the population of children aged 6-23 months

Our analyses included 90.3% of all low-income, 66.0% of all lower-middle and 30.3% of all upper-middle income in the world as of 2016. The numbers of countries with prevalence of 50% or higher were 17 (21.3%) for MDD, 45 (56.2%) for MMF, and only 8 (10.1%) for MAD (Supplementary table 2). Figure 1 shows the ecological analyses with countries as the units. There were direct linear associations between log GDP per capita and MDD (R^2^ = 48.5%), MMF (R^2^ = 28.2%) and MAD (R^2^ = 41.4%). MDD was also correlated to MMF (r = 0.68; p<0.001; data not shown).

**Table 2.** Weighted mean prevalence of each complementary feeding indicator by wealth deciles and the slope index of inequality, according to world regions

| World region | Poorest | D2 | D3 | D4 | D5 | D6 | D7 | D8 | D9 | Wealthiest | SII | 95% CI |
| --- | --- | --- | --- | --- | --- | --- | --- | --- | --- | --- | --- | --- |
| Minimum dietary diversity |  |  |  |  |  |  |  |  |  |  |  |  |
| Western & Central Africa | 12.7 | 14.1 | 14.0 | 15.4 | 14.9 | 18.0 | 21.4 | 22.8 | 29.2 | 34.8 | 22.5 | 17.0; 27.9 |
| Eastern & Southern Africa | 14.2 | 16.4 | 17.6 | 19.9 | 21.5 | 24.2 | 26.4 | 29.7 | 36.5 | 45.1 | 30.2 | 24.3; 36.1 |
| Middle East & North Africa | 32.5 | 36.8 | 32.1 | 34.4 | 36.4 | 35.0 | 36.7 | 38.6 | 40.0 | 43.3 | 9.6 | 5.2; 13.9 |
| Eastern Europe & Central Asia | 41.7 | 44.0 | 48.7 | 47.8 | 52.6 | 53.1 | 51.2 | 54.9 | 58.5 | 52.6 | 14.6 | 8.2; 21.0 |
| South Asia | 13.9 | 16.2 | 17.7 | 19.0 | 20.0 | 21.9 | 23.8 | 25.1 | 25.8 | 27.7 | 14.8 | 13.3; 16.3 |
| East Asia & the Pacific | 30.7 | 44.1 | 45.0 | 47.6 | 48.9 | 55.0 | 53.3 | 56.8 | 61.8 | 64.2 | 30.4 | 22.6; 38.2 |
| Latin America & Caribbean | 46.2 | 57.5 | 49.9 | 60.6 | 53.8 | 59.7 | 60.8 | 57.6 | 58.4 | 72.6 | 17.4 | 5.4; 29.4 |
| Minimum meal frequency |  |  |  |  |  |  |  |  |  |  |  |  |
| Western & Central Africa | 37.4 | 38.2 | 38.6 | 39.0 | 40.0 | 37.5 | 38.9 | 40.0 | 42.3 | 45.6 | 6.5 | 2.9; 10.1 |
| Eastern & Southern Africa | 38.8 | 40.6 | 41.5 | 42.5 | 44.3 | 48.0 | 43.9 | 49.2 | 52.9 | 56.2 | 17.4 | 13.8; 21.0 |
| Middle East & North Africa | 57.1 | 59.0 | 59.0 | 59.9 | 62.4 | 64.1 | 63.7 | 63.1 | 69.6 | 74.1 | 15.7 | 10.2; 21.3 |
| Eastern Europe & Central Asia | 66.9 | 62.3 | 67.7 | 67.6 | 68.7 | 70.5 | 71.1 | 72.5 | 72.2 | 71.6 | 8.9 | 4.6; 13.2 |
| South Asia | 36.8 | 39.4 | 41.3 | 40.8 | 43.3 | 43.2 | 44.9 | 46.4 | 49.0 | 51.2 | 14.2 | 12.3; 16.1 |
| East Asia & the Pacific | 67.0 | 67.1 | 66.7 | 73.1 | 72.0 | 74.3 | 74.2 | 72.1 | 78.8 | 79.3 | 13.6 | 10.0; 17.3 |
| Latin America & Caribbean | 62.0 | 66.7 | 64.6 | 71.7 | 73.1 | 77.8 | 72.7 | 82.0 | 78.7 | 79.9 | 20.5 | 16.1; 24.9 |
| Minimum acceptable diet |  |  |  |  |  |  |  |  |  |  |  |  |
| Western & Central Africa | 6.4 | 7.5 | 7.1 | 7.8 | 7.0 | 8.3 | 10.3 | 11.1 | 14.0 | 17.7 | 10.8 | 7.3; 14.3 |
| Eastern & Southern Africa | 7.0 | 8.5 | 9.5 | 11.4 | 11.9 | 13.6 | 13.6 | 15.9 | 21.0 | 27.6 | 19.2 | 14.4; 23.9 |
| Middle East & North Africa | 20.4 | 26.1 | 22.0 | 24.8 | 25.3 | 24.5 | 24.6 | 27.8 | 29.1 | 33.8 | 10.2 | 5.0; 15.4 |
| Eastern Europe & Central Asia | 30.5 | 29.3 | 34.8 | 37.2 | 41.0 | 38.2 | 40.8 | 43.0 | 41.8 | 42.8 | 15.0 | 10.7; 19.2 |
| South Asia | 7.9 | 8.8 | 10.4 | 10.4 | 11.7 | 13.0 | 12.8 | 14.5 | 16.2 | 17.5 | 10.1 | 9.4; 10.9 |
| East Asia & the Pacific | 23.5 | 32.1 | 32.4 | 36.6 | 37.9 | 44.2 | 42.7 | 43.6 | 51.3 | 53.6 | 29.1 | 24.1; 34.2 |
| Latin America & Caribbean | 33.1 | 41.7 | 37.7 | 43.6 | 41.5 | 48.6 | 47.1 | 48.9 | 51.2 | 59.0 | 22.5 | 16.0; 28.9 |

**NOTE:** All estimates were weighted by the size of the population of children aged 6-23 months

**Figure 1:**
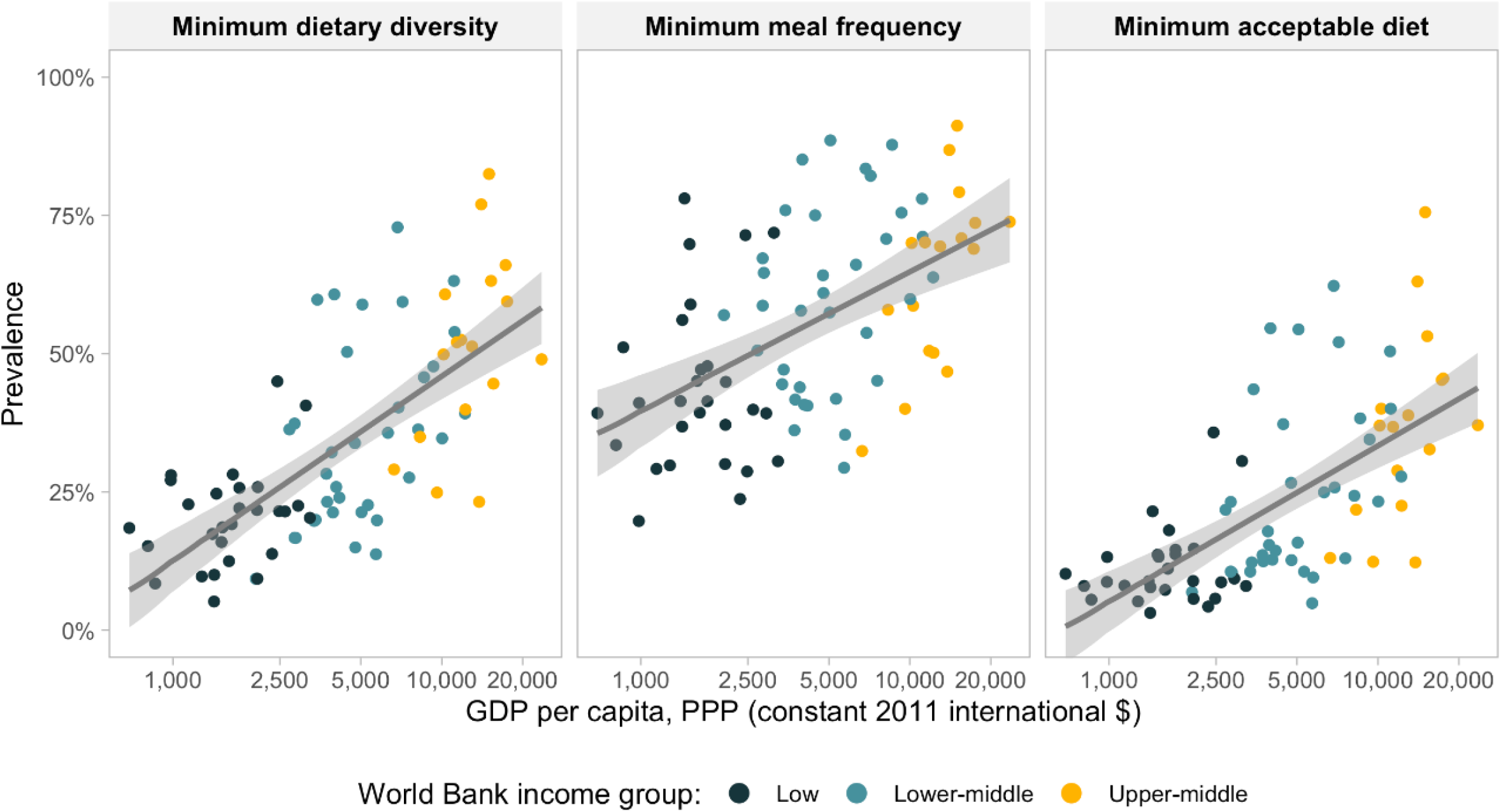
Country-level scatter diagrams of complementary feeding indicators according to per capita GDP.

Table 1 shows prevalence of the three complementary feeding indicators by sex of the child and regions of the world. In all regions, MMF showed higher prevalence than MDD, whereas the lowest prevalence was observed for MAD. The Western & Central Africa was the region with the lowest mean of the weighted prevalence for the three indicators, followed by South Asia and by Eastern and Southern Africa. The highest mean prevalence levels for MDD and MAD were observed in Latin America & Caribbean, and for MMF in East Asia & the Pacific, closely followed by Latin America & Caribbean. No consistent sex differences were observed in the analyses.

Results by wealth decile and region are shown in Table 2. The wealthiest children presented the highest mean values in all regions for the three indicators, and with a couple of minor exceptions the lowest values were observed in the poorest decile. All SII values were positive and significantly different from zero, indicating pro-rich inequality. MDD was more unequally distributed than MFF in six of the seven world regions. The ranking of regions according to inequality varied by indicator, with Latin America & Caribbean being the most unequal region in terms of MMF, whereas MDD was most unequal in East Asia & Pacific and in Eastern & Southern Africa. For MAD, three regions have similar levels of inequality around 10 p.p. (i.e., South Asia, Middle East & North Africa and Western & Central Africa), whereas East Asia & the Pacific presented the highest inequality magnitude at 29.1 p.p.. These results are presented in graphical form in Supplementary Figure 1. Inequality in dietary diversity is being mainly driven by the wealthiest groups in Sub-Saharan Africa and in Latin America & Caribbean, a pattern that has been described as “top inequality”. In contrast, in East Asia & Pacific it is the poorest decile that has markedly lower prevalence than the rest of the population, a “bottom inequality” pattern. When countries are pooled according to World Bank groups, top inequality for dietary diversity is particularly evident in low-income countries. Inequality patterns for meal frequency are not as evident (Supplementary Figure 2).

**Figure 2.**
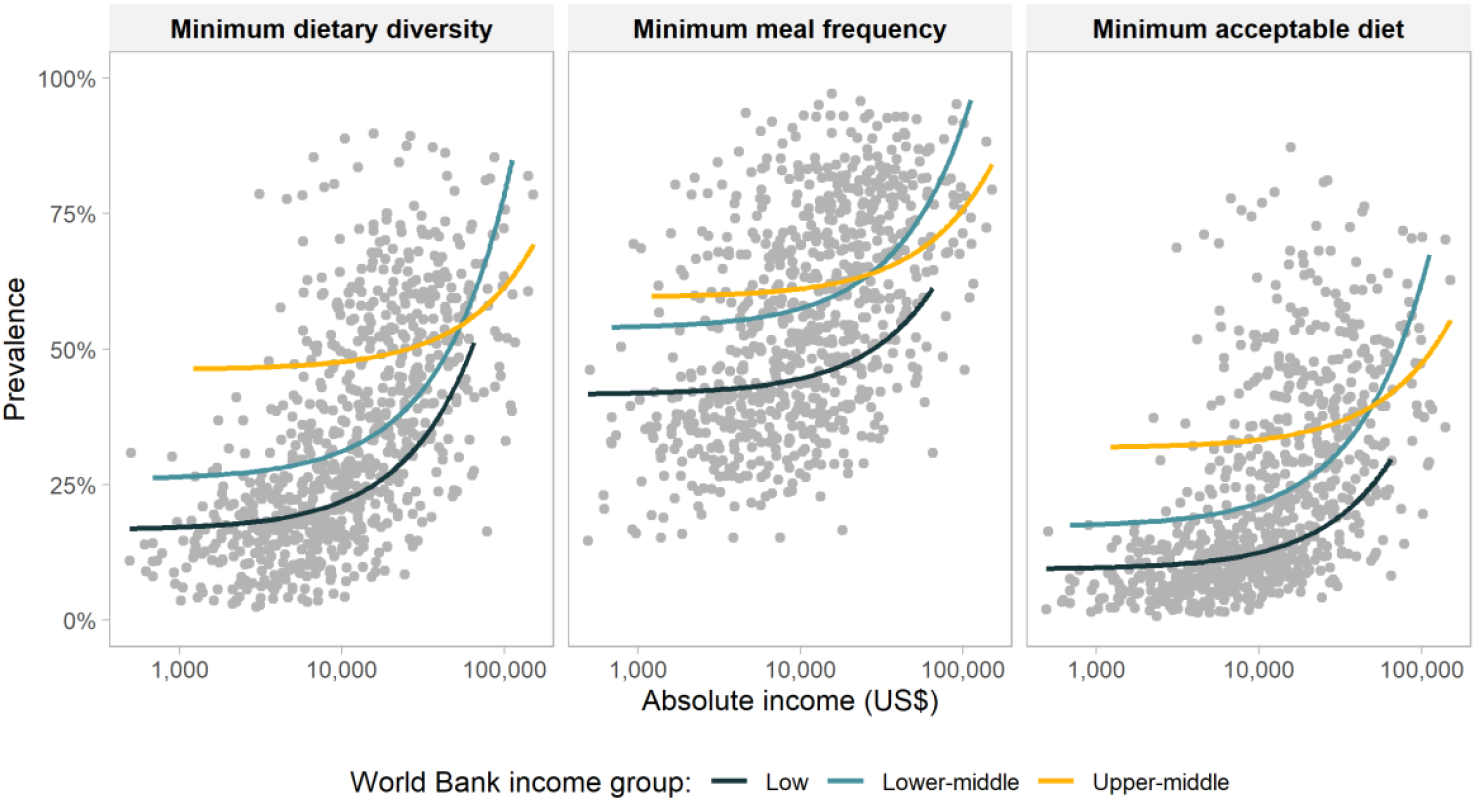
Absolute income and complementary feeding indicators, by World Bank income group. Each dot represents one wealth decile within the 80 countries.

More detailed results by country are presented in Supplementary Tables 3-5 and Supplementary figures 3-9. Most countries showed pro-rich patterns for the three dietary indicators, and out of the 240 analyses performed only two countries presented significant pro-poor patterns (Kiribati and Guinea Bissau) for meal frequency.

**Table 3.** Weighted mean prevalence of consumption of food groups in the poorest and wealthiest deciles, and summary indices of inequality.

| **Food group** | **Poorest**<br>**decile (%)** | **Wealthiest**<br>**decile (%)** | **SII**<br>**(p.p.)** | **95% CI** |
| --- | --- | --- | --- | --- |
| Breastmilk | 36.1 | 28.6 | -8.0 | -9.4; -6.6 |
| Cereal and grains | 33.2 | 33.4 | 0.5 | -0.1; 1.0 |
| Legumes and nuts | 8.6 | 9.4 | 0.9 | 0.1; 1.7 |
| Dairy products | 17.1 | 30.3 | 13.7 | 11.6; 15.7 |
| Flesh foods | 10.7 | 17.2 | 6.6 | 5.9; 7.3 |
| Eggs | 6.9 | 12.8 | 5.8 | 5.1; 6.6 |
| Vitamin A rich fruits and vegetables | 18.5 | 20.2 | 1.2 | 0.5; 1.9 |
| Other fruits and vegetables | 8.7 | 15.1 | 6.4 | 5.6; 7.2 |

SII: slope index of inequality; p.p.: percentage points

**Figure 3.**
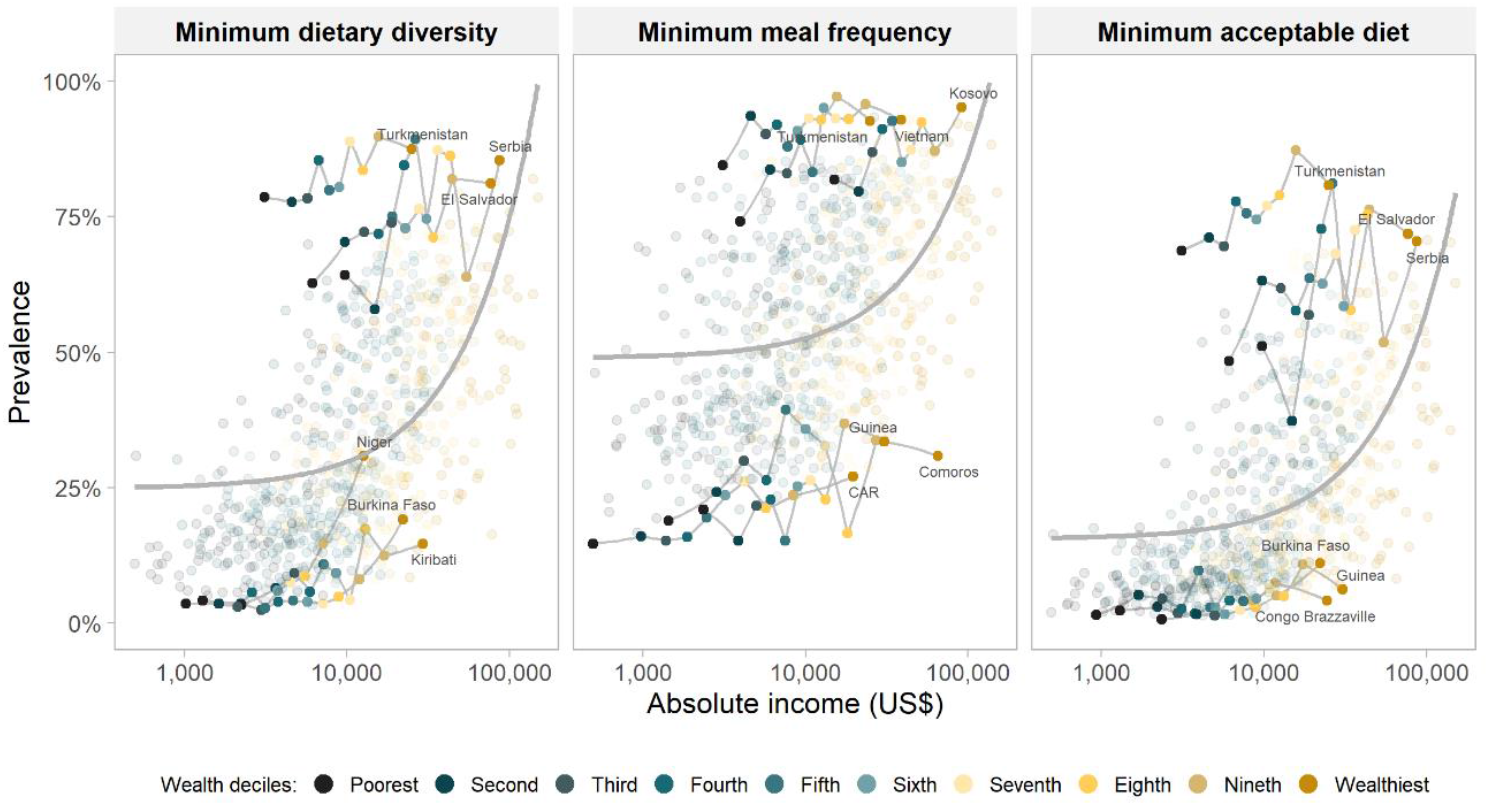
Absolute income and the three top and bottom countries according to prevalence of complementary feeding indicators, by World Bank income group. Each dot represents one wealth decile within the 80 countries.

The sharp inequalities in dietary diversity led us to inspect the role of each of the eight food groups included in this indicator. Breastmilk was the only food with a pro-poor distribution. Inequalities in the consumption of cereal and grains, legumes and nuts, and vitamin A rich fruits and vegetables were small, but for the other four food groups, particularly dairy products, tended to be wide (Table 3). Additional results of food groups by world regions are presented in Supplementary table 6.

In the last set of analyses, the three complementary feeding indicators were plotted against absolute income. In Figure 2, the points represent the 800 deciles in all countries included in the analyses, and the lines are fractional polynomials for the three World Bank country income groups. For the same level of absolute income household income, complementary feeding indicators tend to be highest in upper-middle-income countries, intermediate in lower-middle and in lowest in low-income countries, although at the upper end of scale the patterns are similar in all middle-income countries. These results suggest that country characteristics may be driving complementary feeding patterns, beyond household income levels.

To further analyze these patterns, Figure 3 was derived from Figure 2 to show the top and bottom three countries according to national prevalence of each complementary feeding indicator. Deciles with similar levels of household income show much higher prevalence of dietary indicators in well-performing countries, compare to countries with poor performance. In the latter, even relatively wealthy households show poor child diets. The most marked differences are observed for dietary diversity.

## DISCUSSION

Our analyses add to the literature by presenting the first report on the three internationally recommended indicators of complementary feeding for young children, using the 2018 definition. The main difference between the previous definition from 2007 and the current definition refers to how breastfeeding was counted in terms of dietary diversity, as well as regarding how solid and semi-solid foods are counted regarding meal frequency in non-breastfed children as meal frequency. As a consequence of changes in these two indicators, the acceptable diet variable also changed.

Our pooled results showed that, across the 80 countries studied, only one in four children had diets that were sufficiently diverse, and one in two were fed the number of recommended meals per day. Being a combination of diversity and frequency, minimum acceptable diets were available to only one in six children. Earlier multicountry analyses using the 2007 indicators^13^ or a combination of 2007 and 2018 indicators^15^ had also shown that minimum dietary diversity had lower prevalence than minimum feeding frequency.

We found that East Asia and Pacific was the region with the highest mean MMF, while the Latin America and Caribbean region had the best performance for MDD and MAD. In contrast, the earlier analyses by White et al^13^ showed that East Asia and Pacific had the highest values for the three indicators. These differences may be due to changes in the definition of the indicators, and to the fact that the number of countries in the analyses varied between the two studies. In both studies the lowest mean of the weighted prevalence for all three complementary feeding indicators were observed in the two Sub-Saharan Africa regions and in South Asia.

There were striking wealth related inequalities, with pro-rich patterns present in between-country and within-country analyses, in all seven world regions. Dietary diversity started to improve when absolute household income exceeded about US$20,000. The analyses by Baye and colleagues also found a direct association between GDP and diversity. ^15^

Similar patterns were observed for within-country inequalities, which tended to be wider for diversity than for frequency in five of the seven regions of the world, the exceptions being Latin America & Caribbean and Middle East & North Africa. In contrast to earlier analyses relying on wealth quintiles, our results by wealth decile were able to document socioeconomic gradients with greater granularity, while also confirming earlier reports of wider inequalities for dietary diversity than for frequency at global and regional level.^13,15^

When we analyzed each of the eight food groups, the widest inequalities were observed for consumption of animal-source foods, mainly caused by dairy products consumption, followed by flesh foods and eggs, and for consumption of fruits and vegetables other than those rich in vitamin A. Such inequalities were likely due to the high cost of these foodstuffs. There were virtually no socioeconomic differences in consumption of cereals and grains, legumes and nuts, and vitamin A rich fruits and vegetables. The only food group with higher consumption among children from poor families was breastmilk, a finding that is in accordance with the literature.^26^

The limitations of our study include the fact that indicators are derived from 24-recall by the survey respondents rather than actual observation and measurement of feeding patterns. Although it is possible that respondents may have overreported on the types of foodstuffs and frequencies of feeds, the fact that prevalence of adequate complementary feeding was low suggests that poor diets are a major problem, particularly in low-income countries. Another limitation is that no data were available for 60 of all 140 LMICs ^24^; these included several upper-middle income countries such as China and Brazil, where standardized surveys have not been conducted in recent years.

Among the strengths of our analyses, this is the first comprehensive study that complied with the new definitions of complementary feeding indicators in LMICs, while also addressing both relative and absolute inequalities using different socioeconomic indicators and with greater granularity – wealth deciles rather than quintiles – than previously reported studies. In addition, we explored socioeconomic inequalities according to food groups.

Inadequate complementary feeding practices are a major determinants of poor child growth and development^1–4^. Regular monitoring of dietary adequacy is an essential component for tracking progress towards the health and nutrition-related Sustainable Development Goals.

## Data Availability

The dataset is publicly available.

https://dhsprogram.com/

https://mics.unicef.org/surveys

