## Supplementary tables and figures for "Complementary feeding practices in 80 low- and middle-income countries: prevalence and socioeconomic inequalities in dietary diversity, meal frequency and dietary adequacy"

**Supplementary Table 1.** Countries and surveys included in the complementary feeding inequality analyses. Sources: DHS, MICS and ENSANUT, 2010-2019.

| **Country** | **ISO code** | **Survey and year** | **UNICEF regions** | **Income group** | **Number of children aged 6–23 months** |
|---|---|---|---|---|---|
| Afghanistan | AFG | DHS 2015 | South Asia | LI | 8078 |
| Albania | ALB | DHS 2017 | Eastern Europe & Central Asia | UMI | 766 |
| Angola | AGO | DHS 2015 | Eastern & Southern Africa | UMI | 4009 |
| Armenia | ARM | DHS 2015 | Eastern Europe & Central Asia | LMI | 499 |
| Bangladesh | BGD | MICS 2019 | South Asia | LMI | 6691 |
| Benin | BEN | DHS 2017 | Western & Central Africa | LI | 3884 |
| Burkina Faso | BFA | DHS 2010 | Western & Central Africa | LI | 4148 |
| Burundi | BDI | DHS 2016 | Eastern & Southern Africa | LI | 3858 |
| CAR | CAF | MICS 2010 | Western & Central Africa | LI | 3266 |
| Cambodia | KHM | DHS 2014 | East Asia & the Pacific | LI | 2127 |
| Cameroon | CMR | DHS 2018 | Western & Central Africa | LMI | 2576 |
| Chad | TCD | DHS 2014 | Western & Central Africa | LI | 4403 |
| Comoros | COM | DHS 2012 | Eastern & Southern Africa | LI | 869 |
| Congo Brazzaville | COG | MICS 2014 | Western & Central Africa | LMI | 2765 |
| Congo Democratic Republic | COD | MICS 2017 | Western & Central Africa | LI | 6499 |
| Cote dIvoire | CIV | MICS 2016 | Western & Central Africa | LMI | 2668 |
| Dominican Republic | DOM | MICS 2014 | Latin America & Caribbean | UMI | 6235 |
| Ecuador | ECU | DHS 2012 | Latin America & Caribbean | UMI | 3193 |
| Egypt | EGY | DHS 2014 | Middle East & North Africa | LMI | 4834 |
| El Salvador | SLV | MICS 2014 | Latin America & Caribbean | LMI | 2266 |
| Eswatini | SWZ | MICS 2014 | Eastern & Southern Africa | LMI | 789 |
| Ethiopia | ETH | DHS 2016 | Eastern & Southern Africa | LI | 2822 |
| Gambia | GMB | MICS 2018 | Western & Central Africa | LI | 2724 |
| Georgia | GEO | MICS 2018 | Eastern Europe & Central Asia | UMI | 701 |
| Ghana | GHA | MICS 2017 | Western & Central Africa | LMI | 2585 |
| Guatemala | GTM | DHS 2014 | Latin America & Caribbean | LMI | 3509 |
| Guinea | GIN | DHS 2018 | Western & Central Africa | LI | 1909 |
| Guinea Bissau | GNB | MICS 2014 | Western & Central Africa | LI | 2268 |
| Guyana | GUY | MICS 2014 | Latin America & Caribbean | LMI | 1034 |
| Haiti | HTI | DHS 2016 | Latin America & Caribbean | LI | 1652 |
| Honduras | HND | DHS 2011 | Latin America & Caribbean | LMI | 3237 |
| India | IND | DHS 2015 | South Asia | LMI | 71762 |
| Indonesia | IDN | DHS 2017 | East Asia & the Pacific | LMI | 5033 |
| Iraq | IRQ | MICS 2018 | Middle East & North Africa | UMI | 4786 |
| Jordan | JOR | DHS 2017 | Middle East & North Africa | UMI | 2680 |
| Kazakhstan | KAZ | MICS 2015 | Eastern Europe & Central Asia | UMI | 1632 |
| Kenya | KEN | DHS 2014 | Eastern & Southern Africa | LMI | 2809 |
| Kiribati | KIR | MICS 2018 | East Asia & the Pacific | LMI | 669 |
| Kosovo | XKX | MICS 2013 | Eastern Europe & Central Asia | LMI | 490 |
| Kyrgyzstan | KGZ | MICS 2018 | Eastern Europe & Central Asia | LMI | 992 |
| Lao | LAO | MICS 2017 | East Asia & the Pacific | LMI | 3428 |
| Lesotho | LSO | MICS 2018 | Eastern & Southern Africa | LMI | 1009 |
| Liberia | LBR | DHS 2013 | Western & Central Africa | LI | 2155 |
| Madagascar | MDG | MICS 2018 | Eastern & Southern Africa | LI | 3859 |

| Country | ISO code | Survey and year | UNICEF regions | Income group | Number of children aged 6–23 months |
|---|---|---|---|---|---|
| Malawi | MWI | DHS 2015 | Eastern & Southern Africa | LI | 4747 |
| Mali | MLI | DHS 2018 | Western & Central Africa | LI | 2713 |
| Mauritania | MRT | MICS 2015 | Western & Central Africa | LMI | 3184 |
| Mexico | MEX | MICS 2015 | Latin America & Caribbean | UMI | 2311 |
| Mongolia | MNG | MICS 2018 | East Asia & the Pacific | LMI | 1674 |
| Montenegro | MNE | MICS 2018 | Eastern Europe & Central Asia | UMI | 332 |
| Mozambique | MOZ | DHS 2011 | Eastern & Southern Africa | LI | 3280 |
| Myanmar | MMR | DHS 2015 | East Asia & the Pacific | LMI | 1339 |
| Namibia | NAM | DHS 2013 | Eastern & Southern Africa | UMI | 1303 |
| Nepal | NPL | DHS 2016 | South Asia | LI | 1463 |
| Niger | NER | DHS 2012 | Western & Central Africa | LI | 3260 |
| Nigeria | NGA | DHS 2018 | Western & Central Africa | LMI | 8883 |
| Pakistan | PAK | DHS 2017 | South Asia | LMI | 2566 |
| Papua New Guinea | PNG | DHS 2016 | East Asia & the Pacific | LMI | 2537 |
| Paraguay | PRY | MICS 2016 | Latin America & Caribbean | UMI | 1443 |
| Rwanda | RWA | DHS 2014 | Eastern & Southern Africa | LI | 2354 |
| São Tome and Principe | STP | MICS 2014 | Western & Central Africa | LMI | 571 |
| Senegal | SEN | DHS 2017 | Western & Central Africa | LI | 3489 |
| Serbia | SRB | MICS 2014 | Eastern Europe & Central Asia | UMI | 795 |
| Sierra Leone | SLE | MICS 2017 | Western & Central Africa | LI | 3411 |
| South Africa | ZAF | DHS 2016 | Eastern & Southern Africa | UMI | 877 |
| State of Palestine | PSE | MICS 2014 | Middle East & North Africa | LMI | 2326 |
| Sudan | SDN | MICS 2014 | Eastern & Southern Africa | LMI | 4064 |
| Suriname | SUR | MICS 2018 | Latin America & Caribbean | UMI | 1182 |
| Tajikistan | TJK | DHS 2017 | Eastern Europe & Central Asia | LI | 1722 |
| Tanzania | TZA | DHS 2015 | Eastern & Southern Africa | LI | 3020 |
| Thailand | THA | MICS 2015 | East Asia & the Pacific | UMI | 3222 |
| Timor Leste | TLS | DHS 2016 | East Asia & the Pacific | LMI | 1950 |
| Togo | TGO | MICS 2017 | Western & Central Africa | LI | 1461 |
| Tunisia | TUN | MICS 2018 | Middle East & North Africa | LMI | 946 |
| Turkmenistan | TKM | MICS 2015 | Eastern Europe & Central Asia | UMI | 1169 |
| Uganda | UGA | DHS 2016 | Eastern & Southern Africa | LI | 4160 |
| Vietnam | VNM | MICS 2013 | East Asia & the Pacific | LMI | 1118 |
| Yemen | YEM | DHS 2013 | Middle East & North Africa | LMI | 4299 |
| Zambia | ZMB | DHS 2018 | Eastern & Southern Africa | LMI | 2785 |
| Zimbabwe | ZWE | MICS 2019 | Eastern & Southern Africa | LMI | 1737 |

**NOTE:** DHS: Demographic Health Survey; MICS: Multiple Indicator Cluster Survey; ENSANUT: Encuesta Nacional de Salud y Nutrición; ISO: International Organization for Standardization; LI: Low income country; LMI: Lower-middle income country; UMI: Upper-middle income country. CAR: Central African Republic.

**Supplementary Table 2.** Number of countries according to prevalence ranges for the minimum dietary diversity, minimum meal frequency and minimum acceptable diet indicators

| **Prevalence range (%)** | **Minimum dietary diversity N (%)** | **Minimum meal frequency N (%)** | **Minimum acceptable diet N (%)** |
|---|---|---|---|
| < 10 | 5 (6.3%) | 0 (0%) | 19 (23.7%) |
| 10 to 24.9 | 31 (38.7%) | 2 (2.5%) | 34 (42.5%) |
| 25 to 49.9 | 27 (33.7%) | 33 (41.3%) | 19 (23.7%) |
| 50 to 74.9 | 15 (18.8%) | 32 (40.0%) | 7 (8.8%) |
| 75 to 100 | 2 (2.5%) | 13 (16.2%) | 1 (1.3%) |
| TOTAL | 80 (100%) | 80 (100%) | 80 (100%) |

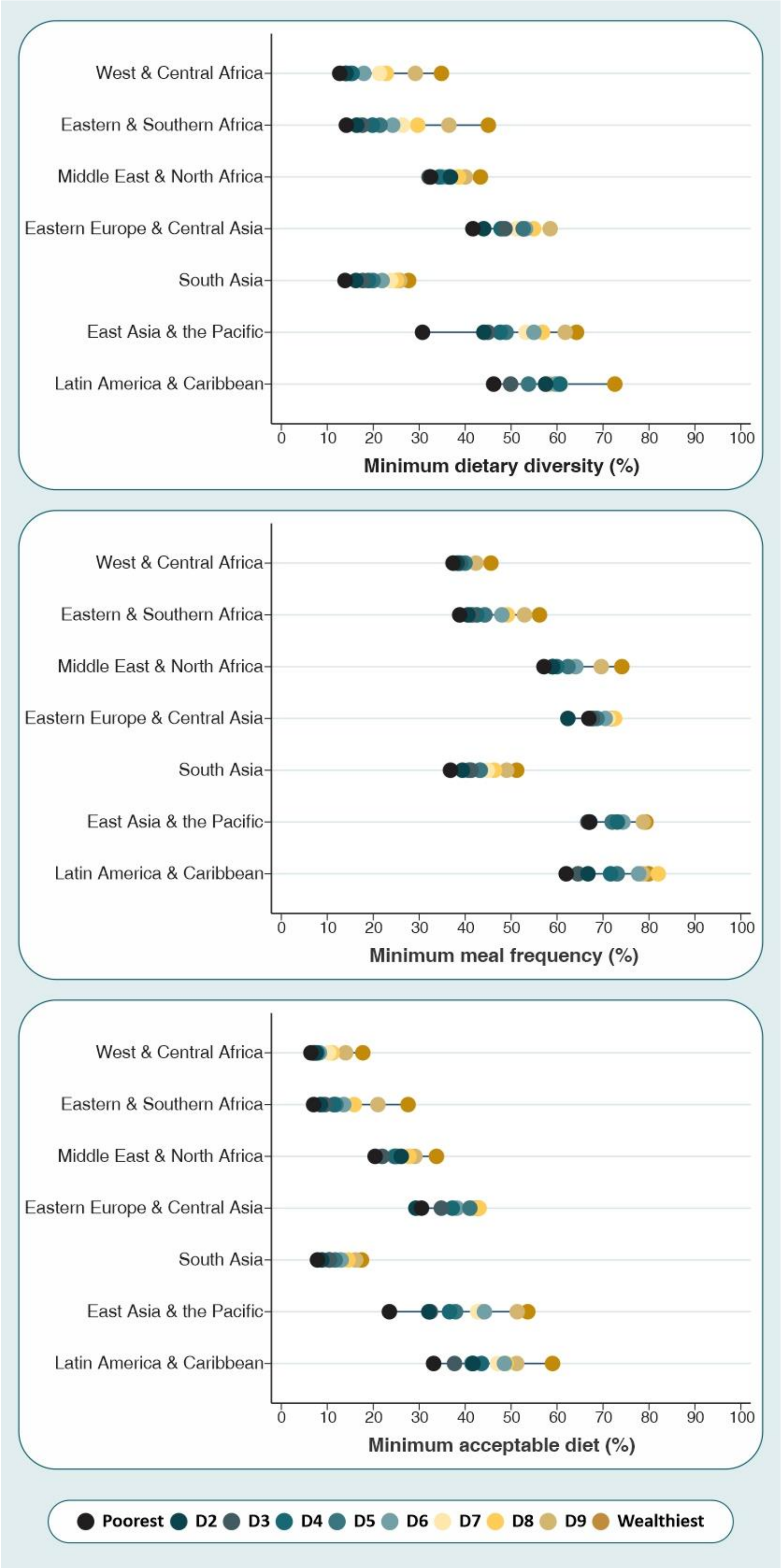


**Supplementary figure 1.** Weighted mean prevalence of dietary indicators by wealth decile, by world region

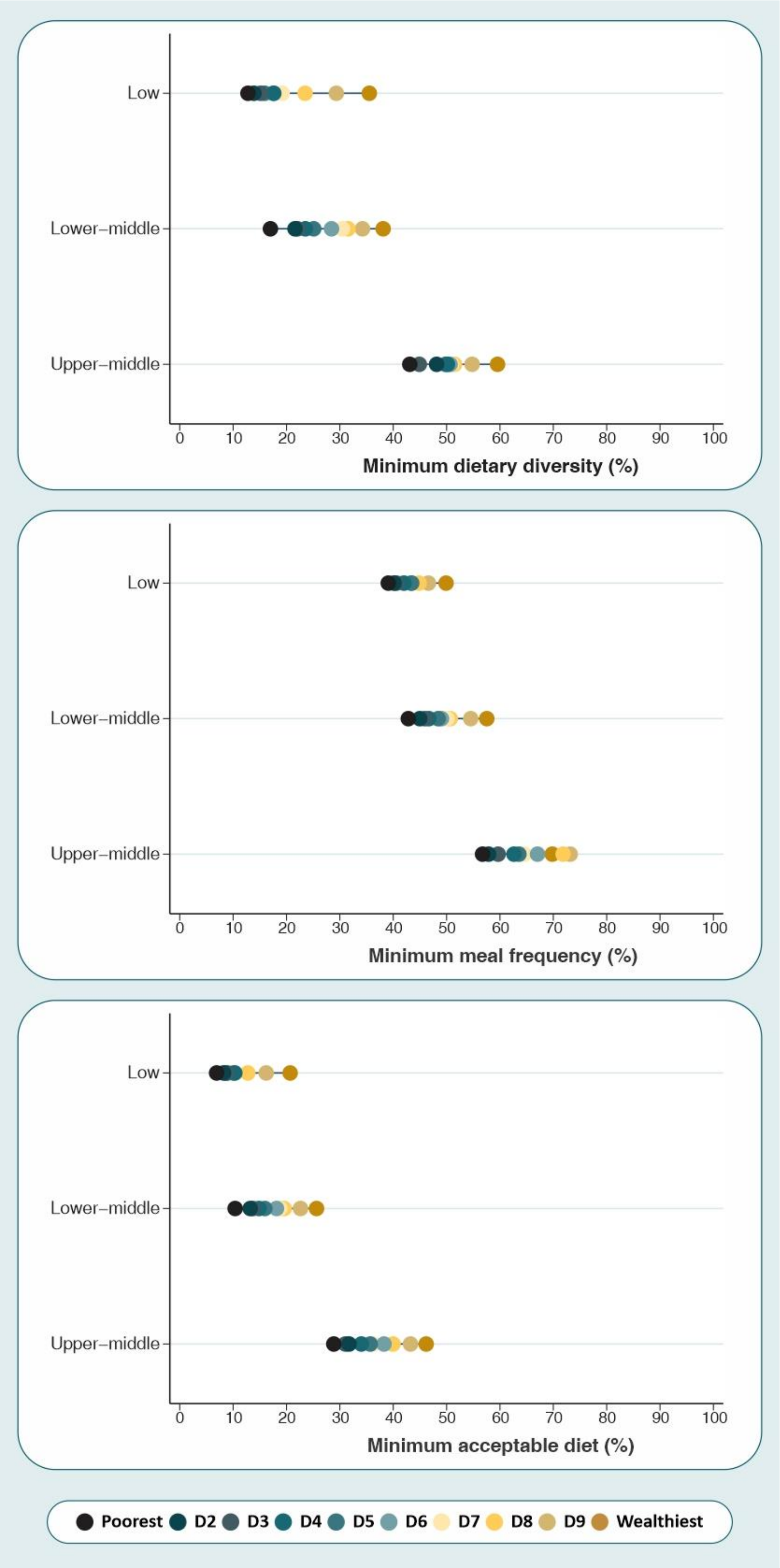


**Supplementary figure 2.** Weighted mean prevalence of dietary indicators by wealth decile, by country income groups

**Supplementary table 3**: Prevalence of minimum dietary diversity by wealth deciles and the magnitude of inequality according to the slope index of inequality

| Country | Year | Poorest | D2 | D3 | D4 | D5 | D6 | D7 | D8 | D9 | Wealthiest | SII | 95% CI |
|---|---|---|---|---|---|---|---|---|---|---|---|---|---|
| | | | | | Western & Central Africa | | | | | | | | |
| Benin | 2017 | 21.2 | 25.5 | 22.2 | 29.6 | 28.2 | 27.9 | 27.8 | 23.7 | 25.7 | 27.3 | 3.4 | -3.6; 10.4 |
| Burkina Faso | 2010 | 4.1 | 3.6 | 2.4 | 3.9 | 4.1 | 3.9 | 3.6 | 4.8 | 8.1 | 19.1 | 8.5 | 4.5; 12.5 |
| CAR | 2010 | 10.9 | 18.0 | 21.0 | 24.7 | 26.5 | 23.6 | 33.3 | 36.1 | 45.5 | 46.0 | 35.3 | 27.6; 42.9 |
| Cote dIvoire | 2016 | 19.5 | 12.8 | 12.0 | 19.5 | 18.5 | 23.1 | 20.7 | 17.1 | 32.9 | 30.0 | 14.4 | 5.7; 23.0 |
| Cameroon | 2018 | 5.7 | 11.7 | 17.8 | 15.9 | 20.4 | 21.3 | 31.3 | 28.3 | 27.3 | 29.5 | 25.5 | 17.3; 33.8 |
| Congo Democratic Republic | 2017 | 10.9 | 12.6 | 14.0 | 17.1 | 10.3 | 11.2 | 13.7 | 19.7 | 24.8 | 23.2 | 11.5 | 5.2; 17.8 |
| Congo Brazzaville | 2014 | 8.8 | 18.3 | 12.6 | 16.9 | 19.2 | 10.1 | 12.0 | 8.8 | 20.8 | 8.4 | -0.0 | -6.1; 6.1 |
| Ghana | 2017 | 18.3 | 18.7 | 24.0 | 12.8 | 24.8 | 23.4 | 21.4 | 20.8 | 44.7 | 51.5 | 29.8 | 21.1; 38.5 |
| Guinea | 2018 | 6.6 | 8.3 | 6.0 | 14.0 | 10.4 | 11.7 | 21.9 | 16.7 | 26.4 | 21.0 | 20.4 | 14.2; 26.6 |
| Gambia | 2018 | 14.6 | 16.9 | 16.2 | 17.1 | 13.3 | 16.2 | 7.9 | 13.6 | 22.1 | 22.8 | 3.0 | -4.6; 10.7 |
| Guinea Bissau | 2014 | 11.2 | 9.1 | 10.6 | 9.7 | 8.5 | 7.4 | 7.2 | 13.5 | 9.1 | 19.0 | 2.1 | -4.2; 8.5 |
| Liberia | 2013 | 10.3 | 7.5 | 9.4 | 3.4 | 9.9 | 8.2 | 14.2 | 12.6 | 10.7 | 13.6 | 5.2 | -2.0; 12.3 |
| Mali | 2018 | 21.4 | 12.9 | 14.3 | 13.0 | 15.0 | 14.8 | 18.3 | 31.2 | 38.4 | 46.3 | 28.0 | 19.0; 37.1 |
| Mauritania | 2015 | 15.3 | 13.0 | 17.6 | 22.9 | 28.0 | 28.9 | 35.3 | 34.8 | 42.8 | 48.4 | 37.9 | 30.2; 45.7 |
| Niger | 2012 | 3.5 | 3.5 | 3.0 | 5.7 | 2.8 | 6.0 | 7.5 | 8.6 | 14.6 | 30.8 | 22.5 | 17.5; 27.5 |
| Nigeria | 2018 | 15.0 | 17.6 | 15.7 | 16.9 | 16.6 | 23.0 | 28.1 | 26.9 | 32.8 | 42.4 | 25.4 | 20.8; 29.9 |
| Senegal | 2017 | 5.5 | 10.9 | 9.2 | 14.0 | 17.3 | 18.8 | 22.3 | 40.2 | 39.9 | 44.4 | 42.2 | 34.5; 49.9 |
| Sierra Leone | 2017 | 13.2 | 14.3 | 15.7 | 14.5 | 14.8 | 19.8 | 16.4 | 21.6 | 27.1 | 25.5 | 13.1 | 6.4; 19.8 |
| Sao Tome and Principe | 2014 | 28.2 | 30.4 | 33.7 | 39.2 | 34.2 | 41.0 | 44.8 | 40.5 | 32.4 | 53.9 | 18.9 | 4.8; 32.9 |
| Chad | 2014 | 5.0 | 4.3 | 4.1 | 6.4 | 5.6 | 8.0 | 10.4 | 15.3 | 14.8 | 26.9 | 19.3 | 14.2; 24.4 |
| Togo | 2017 | 12.3 | 16.6 | 16.0 | 15.6 | 19.8 | 18.2 | 17.0 | 18.4 | 24.2 | 29.4 | 13.2 | 4.1; 22.2 |
| | | | | | Eastern & Southern Africa | | | | | | | | |
| Angola | 2015 | 15.7 | 22.7 | 22.3 | 24.9 | 29.4 | 29.1 | 37.7 | 31.3 | 46.7 | 42.0 | 28.1 | 18.5; 37.7 |
| Burundi | 2016 | 8.0 | 9.7 | 12.7 | 10.5 | 13.9 | 22.5 | 16.5 | 26.1 | 29.3 | 50.9 | 33.2 | 28.1; 38.4 |
| Comoros | 2012 | 17.0 | 21.1 | 16.5 | 23.5 | 15.5 | 27.0 | 22.5 | 26.2 | 19.5 | 27.5 | 8.4 | -5.3; 22.0 |
| Ethiopia | 2016 | 5.7 | 7.3 | 6.2 | 14.0 | 8.3 | 15.6 | 10.2 | 14.8 | 20.9 | 29.6 | 19.9 | 11.5; 28.4 |
| Kenya | 2014 | 15.1 | 22.4 | 26.6 | 25.0 | 33.2 | 34.3 | 45.3 | 55.7 | 53.5 | 63.3 | 50.4 | 42.7; 58.2 |

| Country | Year | Poorest | D2 | D3 | D4 | D5 | D6 | D7 | D8 | D9 | Wealthiest | SII | 95% CI |
|---|---|---|---|---|---|---|---|---|---|---|---|---|---|
| Lesotho | 2018 | 6.0 | 17.5 | 10.4 | 16.9 | 14.6 | 10.4 | 17.7 | 22.7 | 28.5 | 30.2 | 19.8 | 7.7; 31.9 |
| Madagascar | 2018 | 6.7 | 11.1 | 16.9 | 21.9 | 23.2 | 21.5 | 30.6 | 31.5 | 52.6 | 60.8 | 47.7 | 42.4; 53.0 |
| Mozambique | 2011 | 30.8 | 30.2 | 26.5 | 27.4 | 31.7 | 30.7 | 26.9 | 18.0 | 29.5 | 27.7 | -5.1 | -13.0; 2.8 |
| Malawi | 2015 | 13.9 | 14.6 | 16.7 | 22.0 | 19.3 | 23.5 | 23.3 | 26.2 | 36.2 | 47.8 | 28.3 | 22.0; 34.6 |
| Namibia | 2013 | 11.5 | 6.8 | 14.6 | 14.5 | 25.5 | 22.0 | 38.2 | 36.4 | 47.7 | 60.6 | 47.6 | 38.3; 56.8 |
| Rwanda | 2014 | 14.2 | 15.6 | 23.3 | 19.6 | 23.9 | 24.3 | 32.3 | 50.6 | 34.5 | 58.1 | 41.0 | 34.5; 47.4 |
| Sudan | 2014 | 8.6 | 10.0 | 16.1 | 15.7 | 18.8 | 27.4 | 26.7 | 27.0 | 43.9 | 59.4 | 43.7 | 35.6; 51.7 |
| Eswatini | 2014 | 28.1 | 47.7 | 40.6 | 43.8 | 40.7 | 52.4 | 66.6 | 60.1 | 62.2 | 52.0 | 30.6 | 16.6; 44.7 |
| Tanzania | 2015 | 13.5 | 11.8 | 12.3 | 13.6 | 17.3 | 19.8 | 27.9 | 28.3 | 33.9 | 48.7 | 32.8 | 26.5; 39.0 |
| Uganda | 2016 | 18.3 | 21.1 | 25.3 | 24.4 | 25.3 | 22.2 | 23.8 | 30.7 | 31.6 | 36.8 | 16.1 | 9.6; 22.6 |
| South Africa | 2016 | 36.8 | 38.9 | 33.7 | 32.5 | 39.5 | 40.0 | 40.1 | 45.6 | 51.6 | 51.2 | 15.5 | 1.2; 29.9 |
| Zambia | 2018 | 10.8 | 12.1 | 16.7 | 20.2 | 18.0 | 22.1 | 30.7 | 31.5 | 39.0 | 52.8 | 37.3 | 30.1; 44.5 |
| Zimbabwe | 2019 | 6.0 | 8.0 | 7.4 | 8.7 | 15.2 | 16.0 | 16.2 | 29.3 | 26.8 | 44.5 | 36.5 | 28.2; 44.7 |
| Middle East & North Africa | | | | | | | | | | | | | |
| Egypt | 2014 | 35.1 | 39.3 | 29.9 | 34.2 | 36.1 | 33.9 | 34.4 | 33.4 | 33.3 | 38.6 | 0.3 | -5.7; 6.3 |
| Iraq | 2018 | 39.7 | 45.0 | 47.4 | 42.3 | 45.0 | 39.9 | 41.7 | 47.7 | 52.0 | 46.7 | 5.9 | -2.5; 14.4 |
| Jordan | 2017 | 36.8 | 23.9 | 32.2 | 39.1 | 39.0 | 33.4 | 30.8 | 41.0 | 41.1 | 39.6 | 8.8 | -2.6; 20.1 |
| State of Palestine | 2014 | 34.6 | 42.8 | 42.2 | 49.1 | 53.8 | 50.2 | 56.6 | 59.1 | 63.3 | 65.7 | 31.8 | 23.7; 39.9 |
| Tunisia | 2018 | 50.7 | 55.3 | 55.8 | 64.6 | 57.2 | 67.9 | 59.3 | 71.6 | 79.2 | 70.9 | 25.0 | 14.2; 35.7 |
| Yemen | 2013 | 8.3 | 15.4 | 11.5 | 13.0 | 16.9 | 21.7 | 30.1 | 30.9 | 32.2 | 44.5 | 34.3 | 28.0; 40.6 |
| Eastern Europe & Central Asia | | | | | | | | | | | | | |
| Albania | 2017 | 49.5 | 42.5 | 58.7 | 45.1 | 55.4 | 62.7 | 58.4 | 57.1 | 57.7 | 33.1 | 4.4 | -15.3; 24.0 |
| Armenia | 2015 | 38.9 | 28.4 | 34.7 | 30.9 | 43.2 | 34.6 | 34.0 | 36.6 | 40.1 | 43.1 | 10.0 | -8.3; 28.2 |
| Georgia | 2018 | 29.0 | 51.5 | 35.4 | 53.9 | 58.3 | 49.5 | 51.6 | 57.8 | 43.3 | 61.5 | 18.3 | -0.5; 37.1 |
| Kazakhstan | 2015 | 40.1 | 40.0 | 46.6 | 42.6 | 51.0 | 53.6 | 52.5 | 56.2 | 65.8 | 45.0 | 18.6 | 7.8; 29.4 |
| Kyrgyzstan | 2018 | 41.4 | 63.2 | 58.6 | 58.3 | 64.8 | 67.8 | 53.4 | 64.7 | 62.2 | 73.6 | 17.6 | 5.6; 29.7 |
| Montenegro | 2018 | 61.5 | 56.3 | 59.7 | 42.6 | 60.9 | 64.2 | 70.5 | 73.0 | 81.6 | 82.0 | 31.1 | -5.7; 67.9 |
| Serbia | 2014 | 64.2 | 57.9 | 73.8 | 84.5 | 89.4 | 74.6 | 87.2 | 86.2 | 63.9 | 85.4 | 16.0 | -10.0; 42.0 |
| Tajikistan | 2017 | 18.8 | 18.4 | 26.3 | 19.7 | 20.4 | 24.8 | 18.1 | 23.2 | 32.0 | 23.9 | 6.9 | -1.2; 15.0 |

| Country | Year | Poorest | D2 | D3 | D4 | D5 | D6 | D7 | D8 | D9 | Wealthiest | SII | 95% CI |
|---|---|---|---|---|---|---|---|---|---|---|---|---|---|
| Turkmenistan | 2015 | 78.6 | 77.7 | 78.4 | 85.4 | 79.9 | 80.4 | 88.9 | 83.5 | 89.7 | 87.5 | 11.9 | 3.7; 20.1 |
| Kosovo | 2013 | 33.2 | 30.8 | 35.3 | 41.9 | 59.9 | 48.9 | 38.8 | 55.9 | 59.5 | 52.3 | 28.6 | 13.2; 44.0 |
| South Asia | | | | | | | | | | | | | |
| Afghanistan | 2015 | 18.3 | 17.9 | 25.0 | 25.4 | 15.6 | 14.7 | 17.9 | 17.8 | 29.9 | 40.0 | 12.6 | 3.8; 21.4 |
| Bangladesh | 2019 | 22.3 | 21.0 | 25.7 | 28.8 | 28.4 | 34.8 | 37.5 | 42.8 | 44.8 | 52.4 | 33.2 | 28.9; 37.5 |
| India | 2015 | 14.2 | 16.6 | 17.6 | 19.5 | 20.2 | 20.9 | 24.1 | 24.0 | 22.6 | 23.7 | 11.0 | 9.2; 12.9 |
| Nepal | 2016 | 36.8 | 38.6 | 47.2 | 34.2 | 39.5 | 34.8 | 44.2 | 60.5 | 69.7 | 53.0 | 26.8 | 14.9; 38.8 |
| Pakistan | 2017 | 5.1 | 9.6 | 9.7 | 8.8 | 13.5 | 19.7 | 15.0 | 18.0 | 23.7 | 26.7 | 21.9 | 14.4; 29.4 |
| East Asia & the Pacific | | | | | | | | | | | | | |
| Indonesia | 2017 | 30.9 | 48.0 | 47.2 | 48.8 | 50.0 | 59.6 | 55.6 | 60.1 | 67.0 | 71.2 | 35.4 | 29.7; 41.1 |
| Cambodia | 2014 | 22.8 | 34.4 | 32.8 | 39.8 | 38.7 | 36.4 | 42.2 | 55.0 | 51.4 | 60.6 | 35.5 | 26.4; 44.6 |
| Kiribati | 2018 | 3.3 | 6.4 | 9.2 | 5.7 | 10.9 | 9.2 | 4.2 | 17.4 | 12.4 | 14.6 | 10.6 | -0.1; 21.4 |
| Lao | 2017 | 23.8 | 22.5 | 20.6 | 24.6 | 36.5 | 33.9 | 44.8 | 50.4 | 55.7 | 61.8 | 44.0 | 37.7; 50.4 |
| Myanmar | 2015 | 17.1 | 15.7 | 10.2 | 21.1 | 15.7 | 27.7 | 27.2 | 21.3 | 33.2 | 36.3 | 21.3 | 10.4; 32.3 |
| Mongolia | 2018 | 13.3 | 12.8 | 19.2 | 37.3 | 35.7 | 50.1 | 52.2 | 45.6 | 67.2 | 58.0 | 56.4 | 46.3; 66.5 |
| Papua New Guinea | 2016 | 31.2 | 22.0 | 25.6 | 32.9 | 26.4 | 27.9 | 34.4 | 31.9 | 41.5 | 51.3 | 19.4 | 7.8; 31.0 |
| Thailand | 2015 | 69.9 | 57.6 | 61.2 | 66.1 | 75.1 | 66.0 | 54.3 | 63.2 | 55.7 | 61.1 | -5.7 | -20.7; 9.2 |
| Timor Leste | 2016 | 14.2 | 20.2 | 25.6 | 16.4 | 25.9 | 24.9 | 26.0 | 37.6 | 34.6 | 55.7 | 32.1 | 21.0; 43.2 |
| Vietnam | 2013 | 22.9 | 51.4 | 60.3 | 58.2 | 60.5 | 62.4 | 67.8 | 70.1 | 71.6 | 63.6 | 36.0 | 25.4; 46.5 |
| Latin America & Caribbean | | | | | | | | | | | | | |
| Dominican Republic | 2014 | 39.9 | 49.1 | 53.5 | 52.1 | 42.3 | 60.2 | 48.3 | 57.0 | 60.9 | 60.1 | 17.1 | 9.9; 24.4 |
| Ecuador | 2012 | 53.4 | 56.3 | 58.0 | 67.8 | 60.7 | 61.6 | 72.1 | 66.0 | 52.4 | 63.4 | 9.2 | -0.5; 18.9 |
| Guatemala | 2014 | 44.9 | 49.6 | 53.4 | 52.2 | 62.4 | 64.4 | 71.2 | 70.8 | 69.3 | 75.8 | 33.6 | 26.9; 40.3 |
| Guyana | 2014 | 39.6 | 42.9 | 24.2 | 47.1 | 38.0 | 42.1 | 41.8 | 36.3 | 46.9 | 53.3 | 9.5 | -3.3; 22.4 |
| Honduras | 2011 | 41.1 | 57.7 | 60.5 | 60.0 | 57.8 | 61.4 | 73.0 | 68.4 | 68.0 | 72.1 | 27.6 | 20.5; 34.6 |
| Haiti | 2016 | 9.0 | 14.5 | 10.9 | 20.0 | 14.8 | 20.3 | 24.9 | 27.2 | 32.5 | 36.4 | 26.5 | 18.4; 34.7 |
| Mexico | 2015 | 50.5 | 65.0 | 50.1 | 66.8 | 55.5 | 62.9 | 61.4 | 56.1 | 58.1 | 78.5 | 10.5 | -1.0; 22.1 |
| Paraguay | 2016 | 37.5 | 46.5 | 55.6 | 51.5 | 53.4 | 52.7 | 51.5 | 59.8 | 59.3 | 72.3 | 23.4 | 11.9; 35.0 |
| El Salvador | 2014 | 62.8 | 70.3 | 72.2 | 71.8 | 75.0 | 72.9 | 76.5 | 71.2 | 82.0 | 81.2 | 15.3 | 6.6; 24.0 |
| Suriname | 2018 | 10.2 | 18.4 | 18.1 | 28.9 | 41.3 | 23.6 | 26.7 | 23.7 | 31.9 | 16.3 | 16.9 | 5.9; 27.9 |

**Supplementary table 4**: Prevalence of minimum meal frequency by wealth deciles and the magnitude of inequality according to the slope index of inequality

| Country | Year | Poorest | D2 | D3 | D4 | D5 | D6 | D7 | D8 | D9 | Wealthiest | SII | 95% CI |
|---|---|---|---|---|---|---|---|---|---|---|---|---|---|
| Western & Central Africa | | | | | | | | | | | | | |
| Benin | 2017 | 41.2 | 43.5 | 46.1 | 40.1 | 45.2 | 48.8 | 43.8 | 42.9 | 47.5 | 51.7 | 6.7 | -0.4; 13.7 |
| Burkina Faso | 2010 | 35.7 | 36.2 | 37.4 | 35.4 | 34.2 | 32.8 | 37.9 | 37.7 | 38.0 | 47.1 | 5.6 | -0.7; 11.8 |
| CAR | 2010 | 14.6 | 16.0 | 15.2 | 15.9 | 19.4 | 23.6 | 26.2 | 21.2 | 23.5 | 27.1 | 13.8 | 6.9; 20.7 |
| Cote dIvoire | 2016 | 41.0 | 40.9 | 46.3 | 56.6 | 55.2 | 41.1 | 46.0 | 48.3 | 52.0 | 46.2 | 7.5 | -4.9; 19.9 |
| Cameroon | 2018 | 44.8 | 37.1 | 40.5 | 40.9 | 39.7 | 47.4 | 42.9 | 51.1 | 54.2 | 55.1 | 15.0 | 5.6; 24.4 |
| Congo Democratic Republic | 2017 | 34.1 | 33.6 | 31.6 | 35.0 | 37.4 | 31.7 | 31.4 | 34.5 | 34.1 | 30.9 | -1.3 | -10.2; 7.5 |
| Congo Brazzaville | 2014 | 16.7 | 22.4 | 23.9 | 29.0 | 28.5 | 28.7 | 35.5 | 32.7 | 35.1 | 55.8 | 27.5 | 18.5; 36.6 |
| Ghana | 2017 | 27.7 | 36.3 | 42.0 | 37.4 | 38.3 | 37.8 | 47.0 | 39.2 | 50.5 | 52.1 | 20.9 | 11.3; 30.6 |
| Guinea | 2018 | 21.0 | 15.2 | 21.7 | 22.8 | 15.2 | 25.2 | 26.4 | 22.8 | 36.9 | 33.5 | 16.4 | 8.2; 24.6 |
| Gambia | 2018 | 70.9 | 67.5 | 66.9 | 67.7 | 69.2 | 71.7 | 69.9 | 70.5 | 69.0 | 76.5 | 4.5 | -4.8; 13.8 |
| Guinea Bissau | 2014 | 64.6 | 59.3 | 57.3 | 54.2 | 58.9 | 55.8 | 51.4 | 49.2 | 51.4 | 57.5 | -12.1 | -24.1; -0.1 |
| Liberia | 2013 | 24.3 | 21.0 | 31.8 | 27.0 | 25.9 | 33.1 | 31.1 | 30.6 | 42.1 | 39.2 | 16.5 | 4.0; 29 |
| Mali | 2018 | 24.8 | 26.5 | 26.6 | 29.3 | 34.0 | 29.5 | 27.1 | 29.0 | 38.6 | 36.0 | 10.9 | 2.6; 19.2 |
| Mauritania | 2015 | 35.7 | 30.3 | 30.2 | 34.4 | 28.7 | 35.6 | 39.7 | 46.4 | 37.5 | 44.8 | 14.2 | 4.9; 23.5 |
| Niger | 2012 | 40.6 | 48.4 | 45.5 | 55.3 | 53.9 | 49.9 | 57.9 | 54.8 | 49.7 | 53.3 | 9.6 | 2.0; 17.3 |
| Nigeria | 2018 | 42.3 | 42.9 | 43.5 | 39.7 | 41.0 | 37.9 | 39.3 | 40.8 | 42.1 | 50.4 | 1.7 | -4.0; 7.3 |
| Senegal | 2017 | 26.6 | 28.5 | 28.1 | 31.5 | 32.7 | 28.7 | 28.8 | 28.9 | 35.1 | 40.2 | 8.8 | 0.4; 17.3 |
| Sierra Leone | 2017 | 34.3 | 34.5 | 35.5 | 36.0 | 41.1 | 43.0 | 46.7 | 44.0 | 51.7 | 63.9 | 24.4 | 16.7; 32.1 |
| Sao Tome and Principe | 2014 | 55.8 | 49.3 | 45.5 | 62.3 | 47.1 | 72.3 | 60.8 | 60.2 | 73.9 | 68.2 | 22.8 | 5.2; 40.3 |
| Chad | 2014 | 30.0 | 34.6 | 26.5 | 36.1 | 38.0 | 41.1 | 42.1 | 48.3 | 44.2 | 34.0 | 15.5 | 8.3; 22.8 |
| Togo | 2017 | 50.4 | 61.8 | 53.4 | 66.6 | 60.6 | 60.5 | 55.8 | 50.2 | 62.0 | 70.9 | 8.0 | -2.3; 18.3 |
| Eastern & Southern Africa | | | | | | | | | | | | | |
| Angola | 2015 | 30.2 | 26.6 | 23.4 | 23.8 | 28.6 | 32.6 | 30.7 | 42.0 | 51.8 | 48.5 | 24.4 | 16.2; 32.6 |
| Burundi | 2016 | 32.9 | 35.2 | 31.7 | 32.7 | 37.9 | 42.8 | 32.4 | 41.4 | 52.9 | 64.1 | 23.7 | 16.2; 31.3 |
| Comoros | 2012 | 18.9 | 24.2 | 29.9 | 26.4 | 39.4 | 35.8 | 32.7 | 16.6 | 33.8 | 30.9 | 8.6 | -6.8; 24.0 |
| Ethiopia | 2016 | 36.5 | 40.1 | 41.6 | 41.2 | 46.9 | 50.5 | 41.2 | 48.4 | 51.3 | 58.6 | 18.3 | 9.5; 27.1 |
| Kenya | 2014 | 37.8 | 46.9 | 41.5 | 50.3 | 49.7 | 53.0 | 53.3 | 54.0 | 64.9 | 60.1 | 25.5 | 15.6; 35.5 |

| Country | Year | Poorest | D2 | D3 | D4 | D5 | D6 | D7 | D8 | D9 | Wealthiest | SII | 95% CI |
|---|---|---|---|---|---|---|---|---|---|---|---|---|---|
| Lesotho | 2018 | 68.7 | 63.6 | 67.3 | 67.6 | 52.5 | 77.8 | 61.2 | 51.6 | 60.7 | 76.6 | -2.8 | -14.3; 8.7 |
| Madagascar | 2018 | 69.4 | 71.3 | 78.5 | 78.7 | 80.3 | 78.3 | 82.7 | 85.2 | 81.4 | 84.1 | 16.0 | 10.2; 21.8 |
| Mozambique | 2011 | 46.1 | 39.5 | 35.6 | 35.4 | 36.7 | 39.3 | 36.9 | 44.2 | 49.7 | 54.3 | 7.4 | -1.4; 16.2 |
| Malawi | 2015 | 23.1 | 25.6 | 26.5 | 29.3 | 31.5 | 29.5 | 27.9 | 33.7 | 32.8 | 39.4 | 13.5 | 7.3; 19.7 |
| Namibia | 2013 | 19.3 | 31.8 | 38.0 | 42.1 | 41.6 | 36.3 | 35.0 | 46.6 | 58.7 | 72.4 | 33.9 | 22.9; 45.0 |
| Rwanda | 2014 | 37.4 | 37.5 | 44.9 | 46.1 | 39.9 | 49.4 | 53.8 | 57.0 | 52.8 | 62.1 | 25.6 | 18.3; 32.9 |
| Sudan | 2014 | 33.7 | 32.9 | 33.5 | 34.3 | 40.9 | 47.7 | 44.2 | 47.0 | 46.8 | 49.2 | 20.4 | 12.2; 28.7 |
| Eswatini | 2014 | 80.4 | 70.6 | 70.5 | 68.4 | 69.3 | 81.6 | 76.0 | 85.7 | 80.1 | 78.2 | 8.3 | -4.8; 21.4 |
| Tanzania | 2015 | 42.0 | 42.1 | 41.2 | 38.3 | 41.9 | 41.9 | 33.5 | 37.8 | 38.1 | 40.8 | -4.7 | -13.5; 4.1 |
| Uganda | 2016 | 35.6 | 35.9 | 39.8 | 42.3 | 39.8 | 43.7 | 38.6 | 43.6 | 43.9 | 52.6 | 13.7 | 7.4; 19.9 |
| South Africa | 2016 | 38.0 | 45.9 | 49.6 | 49.1 | 39.3 | 51.2 | 53.2 | 59.4 | 68.3 | 62.0 | 25.0 | 12.0; 38.0 |
| Zambia | 2018 | 32.6 | 31.0 | 37.2 | 41.2 | 38.6 | 49.0 | 43.2 | 46.8 | 53.9 | 59.2 | 26.8 | 18.8; 34.7 |
| Zimbabwe | 2019 | 56.8 | 59.3 | 64.8 | 67.5 | 74.6 | 67.6 | 67.6 | 70.8 | 73.9 | 77.5 | 19.9 | 10.9; 28.9 |
| Middle East & North Africa | | | | | | | | | | | | | |
| Egypt | 2014 | 57.1 | 57.7 | 54.3 | 54.5 | 58.4 | 60.3 | 59.4 | 57.4 | 67.2 | 74.3 | 14.2 | 8.0; 20.4 |
| Iraq | 2018 | 63.2 | 62.5 | 69.2 | 73.9 | 73.9 | 68.0 | 71.2 | 73.5 | 78.2 | 77.8 | 14.8 | 7.8; 21.9 |
| Jordan | 2017 | 49.9 | 51.8 | 54.5 | 65.1 | 57.4 | 61.9 | 60.3 | 64.7 | 64.6 | 59.5 | 15.4 | 5.2; 25.6 |
| State of Palestine | 2014 | 69.1 | 74.6 | 76.6 | 75.2 | 68.9 | 73.7 | 77.5 | 80.2 | 77.9 | 81.2 | 9.7 | 2.5; 16.9 |
| Tunisia | 2018 | 66.7 | 80.3 | 83.4 | 77.8 | 71.1 | 80.9 | 80.2 | 76.8 | 80.2 | 78.4 | 3.4 | -7.0; 13.8 |
| Yemen | 2013 | 46.8 | 52.9 | 53.0 | 50.8 | 58.1 | 66.0 | 62.0 | 61.4 | 63.4 | 70.2 | 22.2 | 14.7; 29.8 |
| Eastern Europe & Central Asia | | | | | | | | | | | | | |
| Albania | 2017 | 31.5 | 54.0 | 48.3 | 60.0 | 42.5 | 59.4 | 46.3 | 56.3 | 59.5 | 46.2 | 13.4 | -5.4; 32.2 |
| Armenia | 2015 | 71.5 | 70.7 | 73.8 | 75.7 | 79.1 | 62.5 | 54.4 | 67.6 | 73.5 | 76.6 | -0.8 | -16.3; 14.6 |
| Georgia | 2018 | 59.2 | 70.6 | 69.4 | 62.8 | 77.1 | 80.6 | 74.7 | 55.7 | 69.7 | 79.9 | 9.0 | -7.6; 25.6 |
| Kazakhstan | 2015 | 81.9 | 60.8 | 72.3 | 67.7 | 68.1 | 81.9 | 82.3 | 81.5 | 76.7 | 67.6 | 7.1 | -8.6; 22.7 |
| Kyrgyzstan | 2018 | 61.8 | 70.1 | 70.6 | 75.9 | 77.3 | 76.9 | 80.9 | 84.8 | 83.0 | 89.9 | 24.8 | 13.9; 35.7 |
| Montenegro | 2018 | 52.7 | 65.6 | 76.6 | 47.1 | 77.8 | 79.5 | 63.3 | 68.9 | 67.9 | 88.3 | 28.5 | -1.3; 58.3 |
| Serbia | 2014 | 83.9 | 78.3 | 81.2 | 88.9 | 93.3 | 83.9 | 86.7 | 92.9 | 84.6 | 92.3 | 9.8 | -1.2; 20.7 |
| Tajikistan | 2017 | 36.9 | 34.0 | 41.6 | 41.4 | 43.5 | 33.5 | 35.5 | 39.0 | 42.1 | 46.7 | 4.1 | -5.1; 13.4 |
| Turkmenistan | 2015 | 84.5 | 93.6 | 90.3 | 92.0 | 87.9 | 90.9 | 93.1 | 92.9 | 97.1 | 92.7 | 7.5 | 1.0; 14.0 |

| Country | Year | Poorest | D2 | D3 | D4 | D5 | D6 | D7 | D8 | D9 | Wealthiest | SII | 95% CI |
|---|---|---|---|---|---|---|---|---|---|---|---|---|---|
| Kosovo | 2013 | 81.9 | 79.7 | 86.9 | 91.2 | 92.7 | 85.0 | 87.3 | 92.5 | 87.2 | 95.2 | 10.6 | 0.2; 21.0 |
| South Asia | | | | | | | | | | | | | |
| Afghanistan | 2015 | 49.6 | 47.8 | 51.2 | 51.4 | 48.2 | 40.1 | 47.3 | 48.5 | 45.4 | 49.2 | -3.1 | -12.5; 6.2 |
| Bangladesh | 2019 | 58.6 | 60.4 | 59.0 | 61.5 | 63.3 | 63.9 | 67.4 | 68.1 | 69.2 | 70.4 | 14.0 | 9.4; 18.7 |
| India | 2015 | 30.8 | 32.5 | 33.9 | 33.6 | 35.8 | 34.8 | 36.5 | 36.7 | 41.2 | 43.7 | 11.1 | 9.2; 13.0 |
| Nepal | 2016 | 71.8 | 76.9 | 71.5 | 71.3 | 66.9 | 66.1 | 65.2 | 75.8 | 78.8 | 73.0 | 1.1 | -8.6; 10.7 |
| Pakistan | 2017 | 44.5 | 51.7 | 57.8 | 54.8 | 60.8 | 65.3 | 65.2 | 71.8 | 68.7 | 70.7 | 28.2 | 18.6; 37.8 |
| East Asia & the Pacific | | | | | | | | | | | | | |
| Indonesia | 2017 | 67.3 | 65.2 | 63.7 | 72.9 | 69.4 | 72.1 | 73.5 | 66.7 | 78.4 | 81.5 | 14.3 | 8.8; 19.8 |
| Cambodia | 2014 | 67.9 | 60.6 | 69.4 | 68.6 | 71.9 | 74.6 | 69.9 | 75.6 | 79.8 | 83.5 | 19.1 | 9.2; 29.0 |
| Kiribati | 2018 | 59.8 | 68.0 | 58.7 | 62.8 | 67.9 | 46.5 | 58.0 | 53.6 | 48.6 | 48.0 | -18.3 | -31.5; -5.1 |
| Lao | 2017 | 60.1 | 58.6 | 56.5 | 61.0 | 64.0 | 65.1 | 71.4 | 77.0 | 78.1 | 79.1 | 25.5 | 18.9; 32.1 |
| Myanmar | 2015 | 54.2 | 54.1 | 50.3 | 58.4 | 65.5 | 58.2 | 56.0 | 63.6 | 61.1 | 59.2 | 9.6 | -2.1; 21.3 |
| Mongolia | 2018 | 51.7 | 52.7 | 56.2 | 61.8 | 60.8 | 71.5 | 74.0 | 70.0 | 76.3 | 64.2 | 23.9 | 11.5; 36.2 |
| Papua New Guinea | 2016 | 34.1 | 44.4 | 41.8 | 40.8 | 48.3 | 43.6 | 45.1 | 42.6 | 40.1 | 62.6 | 12.9 | 2.2; 23.5 |
| Thailand | 2015 | 79.4 | 77.0 | 84.0 | 76.4 | 85.3 | 79.4 | 75.7 | 82.5 | 81.6 | 67.6 | -2.6 | -15.4; 10.3 |
| Timor Leste | 2016 | 46.3 | 35.9 | 37.0 | 46.1 | 40.2 | 38.5 | 51.5 | 51.4 | 45.5 | 60.2 | 15.9 | 5.8; 25.9 |
| Vietnam | 2013 | 74.2 | 83.7 | 83.0 | 89.3 | 83.3 | 95.0 | 93.1 | 93.1 | 95.8 | 92.8 | 20.6 | 12.9; 28.3 |
| Latin America & Caribbean | | | | | | | | | | | | | |
| Dominican Republic | 2014 | 57.8 | 65.2 | 71.5 | 68.4 | 67.5 | 76.4 | 71.9 | 71.5 | 81.6 | 69.4 | 16.6 | 10.8; 22.3 |
| Ecuador | 2012 | 49.8 | 51.3 | 56.3 | 63.3 | 51.8 | 59.3 | 66.9 | 67.2 | 58.4 | 71.3 | 19.0 | 7.5; 30.5 |
| Guatemala | 2014 | 76.3 | 81.9 | 76.9 | 80.4 | 83.5 | 82.3 | 81.6 | 89.1 | 87.0 | 91.7 | 13.1 | 8.1; 18.2 |
| Guyana | 2014 | 39.3 | 58.3 | 51.4 | 67.1 | 56.3 | 39.7 | 55.9 | 52.3 | 56.3 | 69.7 | 13.2 | 0.5; 25.9 |
| Honduras | 2011 | 81.6 | 86.5 | 83.3 | 85.2 | 78.5 | 85.3 | 89.9 | 90.1 | 85.6 | 86.9 | 6.2 | 0.8; 11.5 |
| Haiti | 2016 | 20.4 | 34.0 | 26.6 | 38.9 | 29.5 | 43.7 | 38.0 | 54.7 | 63.0 | 79.1 | 45.6 | 37.1; 54.1 |
| Mexico | 2015 | 64.4 | 67.5 | 64.5 | 74.3 | 78.7 | 83.2 | 73.3 | 86.5 | 81.1 | 79.4 | 22.2 | 11.7; 32.8 |
| Paraguay | 2016 | 54.7 | 68.3 | 67.8 | 62.0 | 76.8 | 74.7 | 80.1 | 81.3 | 73.8 | 75.8 | 25.0 | 14.0; 36.0 |
| El Salvador | 2014 | 77.6 | 82.5 | 83.3 | 81.8 | 82.1 | 81.2 | 88.9 | 83.5 | 89.6 | 88.8 | 10.1 | 3.1; 17.2 |
| Suriname | 2018 | 40.7 | 41.0 | 46.2 | 44.2 | 48.9 | 59.7 | 49.1 | 51.0 | 42.8 | 56.0 | 13.2 | -0.7; 27.2 |

**Supplementary table 5**: Prevalence of minimum acceptable diet by wealth deciles and the magnitude of inequality according to the slope index of inequality

| Country | Year | Poorest | D2 | D3 | D4 | D5 | D6 | D7 | D8 | D9 | Wealthiest | SII | 95% CI |
|---|---|---|---|---|---|---|---|---|---|---|---|---|---|
| Western & Central Africa | | | | | | | | | | | | | |
| Benin | 2017 | 14.5 | 16.0 | 12.1 | 17.6 | 15.6 | 13.8 | 15.5 | 11.7 | 14.5 | 17.0 | -0.2 | -5.9; 5.5 |
| Burkina Faso | 2010 | 2.3 | 2.9 | 1.8 | 1.7 | 2.8 | 1.6 | 2.4 | 2.9 | 5.1 | 11.0 | 4.7 | 1.8; 7.6 |
| CAR | 2010 | 1.9 | 6.5 | 5.5 | 7.8 | 8.2 | 7.2 | 11.3 | 10.9 | 14.1 | 21.3 | 15.0 | 9.4; 20.6 |
| Cote dIvoire | 2016 | 13.3 | 7.9 | 8.9 | 14.7 | 12.1 | 9.5 | 13.0 | 7.7 | 23.1 | 12.4 | 5.0 | -2.4; 12.4 |
| Cameroon | 2018 | 3.4 | 5.9 | 8.3 | 8.9 | 12.2 | 11.5 | 14.6 | 15.7 | 15.4 | 15.9 | 14.2 | 8.2; 20.2 |
| Congo Democratic Republic | 2017 | 7.0 | 8.8 | 6.9 | 8.8 | 3.9 | 6.8 | 6.4 | 11.0 | 12.0 | 9.5 | 3.0 | -2.2; 8.3 |
| Congo Brazzaville | 2014 | 1.5 | 5.2 | 4.4 | 2.7 | 9.7 | 2.9 | 8.5 | 4.0 | 7.4 | 4.2 | 3.7 | -0.3; 7.7 |
| Ghana | 2017 | 9.6 | 9.6 | 12.1 | 5.0 | 13.1 | 11.3 | 13.7 | 9.2 | 20.2 | 24.1 | 13.3 | 5.7; 21.0 |
| Guinea | 2018 | 0.7 | 1.6 | 1.4 | 4.2 | 4.0 | 4.5 | 6.4 | 5.0 | 10.8 | 6.2 | 9.1 | 4.8; 13.4 |
| Gambia | 2018 | 12.4 | 14.8 | 13.8 | 14.9 | 11.6 | 11.9 | 5.7 | 11.2 | 20.2 | 21.8 | 3.3 | -3.8; 10.5 |
| Guinea Bissau | 2014 | 10.5 | 7.0 | 7.1 | 7.1 | 7.6 | 6.0 | 5.7 | 9.9 | 6.9 | 12.2 | 0.0 | -5.3; 5.4 |
| Liberia | 2013 | 4.8 | 2.3 | 5.2 | 1.4 | 6.3 | 1.8 | 7.2 | 9.4 | 6.9 | 9.7 | 6.0 | -0.8; 12.7 |
| Mali | 2018 | 7.0 | 4.6 | 6.2 | 4.6 | 6.0 | 6.0 | 6.7 | 13.4 | 18.9 | 19.2 | 14.7 | 9.0; 20.5 |
| Mauritania | 2015 | 7.4 | 6.0 | 7.7 | 13.2 | 8.2 | 12.5 | 21.3 | 19.3 | 18.9 | 23.1 | 19.8 | 12.9; 26.8 |
| Niger | 2012 | 2.0 | 3.4 | 1.7 | 4.7 | 2.0 | 5.0 | 5.6 | 5.5 | 7.6 | 18.7 | 12.5 | 8.9; 16.2 |
| Nigeria | 2018 | 6.7 | 8.2 | 7.9 | 8.1 | 6.9 | 9.4 | 12.8 | 12.4 | 14.6 | 23.6 | 13.2 | 9.4; 16.9 |
| Senegal | 2017 | 3.1 | 5.9 | 4.2 | 5.7 | 9.0 | 7.4 | 8.6 | 14.0 | 12.6 | 14.7 | 12.4 | 7.3; 17.5 |
| Sierra Leone | 2017 | 5.3 | 6.0 | 7.3 | 7.3 | 7.6 | 10.7 | 11.4 | 8.6 | 12.2 | 19.7 | 10.5 | 5.6; 15.3 |
| Sao Tome and Principe | 2014 | 14.3 | 15.5 | 27.5 | 27.4 | 17.3 | 34.0 | 26.9 | 17.5 | 29.0 | 27.5 | 11.6 | -3.4; 26.7 |
| Chad | 2014 | 3.4 | 2.9 | 2.1 | 3.0 | 3.8 | 5.1 | 8.2 | 11.8 | 10.8 | 9.0 | 10.5 | 6.7; 14.2 |
| Togo | 2017 | 9.1 | 14.7 | 12.4 | 12.9 | 11.5 | 14.4 | 6.7 | 12.3 | 17.1 | 22.9 | 7.3 | -0.6; 15.2 |
| Eastern & Southern Africa | | | | | | | | | | | | | |
| Angola | 2015 | 6.5 | 9.7 | 9.0 | 8.8 | 14.6 | 13.6 | 15.5 | 14.2 | 24.6 | 20.5 | 16.1 | 8.8; 23.5 |
| Burundi | 2016 | 3.1 | 4.0 | 5.4 | 4.8 | 8.3 | 13.3 | 7.8 | 13.8 | 18.7 | 33.4 | 24.2 | 19.3; 29.0 |
| Comoros | 2012 | 5.7 | 2.6 | 1.4 | 8.1 | 10.3 | 3.0 | 7.1 | 5.2 | 6.9 | 8.2 | 3.1 | -2.7; 8.9 |
| Ethiopia | 2016 | 2.2 | 3.5 | 3.9 | 9.4 | 6.3 | 9.9 | 3.7 | 6.7 | 10.2 | 21.2 | 12.8 | 6.6; 19.0 |
| Kenya | 2014 | 7.9 | 13.4 | 15.7 | 16.0 | 19.5 | 19.1 | 26.2 | 32.8 | 35.3 | 38.1 | 33.0 | 24.9; 41.1 |
| Lesotho | 2018 | 3.7 | 10.3 | 4.9 | 15.4 | 2.6 | 9.0 | 8.1 | 12.6 | 16.0 | 29.4 | 15.6 | 5.5; 25.7 |

| Region/country | Year | Poorest | D2 | D3 | D4 | D5 | D6 | D7 | D8 | D9 | Wealthiest | SII | 95% CI |
|---|---|---|---|---|---|---|---|---|---|---|---|---|---|
| Madagascar | 2018 | 5.5 | 9.3 | 14.9 | 20.1 | 21.7 | 18.3 | 27.7 | 26.4 | 43.3 | 53.5 | 41.5 | 36.0; 47.0 |
| Mozambique | 2011 | 16.3 | 17.5 | 13.3 | 11.1 | 13.7 | 14.0 | 8.5 | 7.8 | 12.5 | 17.8 | -5.3 | -11.6; 1.0 |
| Malawi | 2015 | 3.3 | 3.9 | 4.7 | 8.3 | 7.9 | 6.3 | 9.2 | 12.2 | 12.2 | 20.8 | 14.8 | 10.9; 18.7 |
| Namibia | 2013 | 2.3 | 3.1 | 7.1 | 6.2 | 11.1 | 11.0 | 13.0 | 24.7 | 29.8 | 35.6 | 32.7 | 23.6; 41.8 |
| Rwanda | 2014 | 7.6 | 7.7 | 14.4 | 13.0 | 12.7 | 18.8 | 23.2 | 33.2 | 20.7 | 40.3 | 31.2 | 25.3; 37.2 |
| Sudan | 2014 | 6.2 | 5.1 | 11.3 | 9.9 | 12.8 | 14.5 | 16.9 | 15.4 | 25.5 | 33.3 | 24.4 | 17.3; 31.5 |
| Eswatini | 2014 | 16.3 | 36.2 | 31.6 | 32.6 | 26.7 | 41.2 | 43.9 | 44.5 | 47.0 | 37.4 | 24.1 | 10.2; 37.9 |
| Tanzania | 2015 | 7.4 | 5.4 | 5.1 | 6.5 | 6.7 | 7.5 | 9.2 | 9.5 | 13.3 | 20.3 | 10.8 | 6.1; 15.6 |
| Uganda | 2016 | 8.6 | 13.0 | 13.7 | 14.5 | 13.8 | 13.1 | 13.8 | 16.0 | 18.8 | 21.6 | 10.6 | 5.8; 15.3 |
| South Africa | 2016 | 17.6 | 16.8 | 14.3 | 18.0 | 18.0 | 27.0 | 21.6 | 29.5 | 38.3 | 38.9 | 22.9 | 9.7; 36.1 |
| Zambia | 2018 | 5.5 | 6.8 | 10.5 | 10.9 | 9.3 | 12.3 | 15.7 | 13.8 | 22.2 | 28.8 | 19.8 | 14.1; 25.6 |
| Zimbabwe | 2019 | 4.0 | 5.0 | 6.4 | 7.8 | 8.4 | 9.6 | 14.8 | 16.0 | 15.1 | 25.8 | 20.4 | 13.5; 27.3 |
| Middle East & North Africa | | | | | | | | | | | | | |
| Egypt | 2014 | 21.2 | 28.8 | 18.3 | 24.1 | 24.3 | 21.4 | 22.2 | 22.8 | 23.1 | 29.2 | 2.2 | -3.2; 7.5 |
| Iraq | 2018 | 26.3 | 29.1 | 34.6 | 30.7 | 33.7 | 29.6 | 30.5 | 36.2 | 39.5 | 39.6 | 11.1 | 3.5; 18.7 |
| Jordan | 2017 | 18.0 | 11.1 | 22.2 | 30.8 | 20.6 | 19.4 | 18.0 | 32.4 | 29.4 | 28.5 | 13.1 | 3.4; 22.9 |
| State of Palestine | 2014 | 24.5 | 30.3 | 30.5 | 33.7 | 35.5 | 37.6 | 40.5 | 49.2 | 50.1 | 54.2 | 30.3 | 22.7; 37.9 |
| Tunisia | 2018 | 36.2 | 45.9 | 46.5 | 54.9 | 45.1 | 60.6 | 42.7 | 53.3 | 62.9 | 56.2 | 17.3 | 4.9; 29.7 |
| Yemen | 2013 | 6.1 | 11.5 | 9.4 | 8.5 | 12.0 | 18.4 | 19.1 | 21.3 | 22.6 | 33.4 | 24.5 | 18.5; 30.4 |
| Eastern Europe & Central Asia | | | | | | | | | | | | | |
| Albania | 2017 | 14.1 | 29.8 | 24.7 | 25.0 | 28.1 | 38.9 | 35.4 | 30.1 | 39.5 | 19.6 | 14.0 | -3.0; 31.1 |
| Armenia | 2015 | 29.4 | 14.8 | 21.8 | 23.1 | 33.2 | 21.2 | 12.2 | 26.0 | 28.8 | 33.0 | 9.9 | -6.8; 26.6 |
| Georgia | 2018 | 20.1 | 41.4 | 29.1 | 29.4 | 40.0 | 39.0 | 40.8 | 39.6 | 32.2 | 53.9 | 19.2 | 0.8; 37.7 |
| Kazakhstan | 2015 | 32.2 | 24.7 | 31.9 | 34.2 | 38.5 | 40.1 | 46.2 | 46.2 | 42.4 | 38.1 | 18.2 | 6.7; 29.6 |
| Kyrgyzstan | 2018 | 28.1 | 41.7 | 43.9 | 45.2 | 48.0 | 43.3 | 43.4 | 47.6 | 44.8 | 60.9 | 17.7 | 4.7; 30.7 |
| Montenegro | 2018 | 24.3 | 29.8 | 40.1 | 35.8 | 34.2 | 49.4 | 51.7 | 51.0 | 62.1 | 70.3 | 49.7 | 29.0; 70.3 |
| Serbia | 2014 | 51.1 | 37.3 | 56.9 | 72.8 | 81.2 | 58.5 | 72.5 | 75.4 | 51.8 | 70.4 | 19.3 | -4.1; 42.7 |
| Tajikistan | 2017 | 6.2 | 3.3 | 12.4 | 9.0 | 11.0 | 8.3 | 7.9 | 10.4 | 13.5 | 11.0 | 5.3 | -0.1; 10.7 |
| Turkmenistan | 2015 | 68.8 | 71.1 | 69.5 | 77.9 | 75.7 | 74.4 | 77.0 | 79.0 | 87.3 | 80.8 | 16.1 | 7.2; 24.9 |
| Kosovo | 2013 | 29.3 | 26.7 | 32.2 | 35.3 | 53.1 | 36.6 | 27.4 | 45.4 | 48.4 | 47.4 | 20.8 | 5.8; 35.8 |

| Region/country | Year | Poorest | D2 | D3 | D4 | D5 | D6 | D7 | D8 | D9 | Wealthiest | SII | 95% CI |
|---|---|---|---|---|---|---|---|---|---|---|---|---|---|
| South Asia | | | | | | | | | | | | | |
| Afghanistan | 2015 | 9.7 | 11.3 | 19.2 | 17.4 | 9.0 | 8.7 | 8.5 | 11.4 | 19.1 | 24.1 | 6.7 | -0.6; 14.0 |
| Bangladesh | 2019 | 17.3 | 16.7 | 20.1 | 22.7 | 20.8 | 25.2 | 29.3 | 34.1 | 36.5 | 43.9 | 28.4 | 24.3; 32.6 |
| India | 2015 | 6.8 | 7.6 | 8.8 | 8.8 | 10.4 | 10.4 | 10.6 | 11.1 | 11.8 | 11.8 | 5.7 | 4.4; 7.0 |
| Nepal | 2016 | 29.5 | 32.1 | 39.6 | 27.6 | 29.9 | 27.4 | 33.8 | 48.7 | 56.7 | 35.6 | 18.8 | 6.5; 31.2 |
| Pakistan | 2017 | 5.0 | 7.1 | 7.5 | 7.9 | 11.1 | 17.1 | 11.9 | 15.8 | 19.3 | 24.1 | 19.3 | 11.9; 26.7 |
| East Asia & the Pacific | | | | | | | | | | | | | |
| Indonesia | 2017 | 23.6 | 31.7 | 30.3 | 35.5 | 36.7 | 46.1 | 41.7 | 40.8 | 54.6 | 58.7 | 33.2 | 27.2; 39.2 |
| Cambodia | 2014 | 17.8 | 20.4 | 24.3 | 28.3 | 24.1 | 27.1 | 28.8 | 42.7 | 45.0 | 53.2 | 35.5 | 26.1; 44.8 |
| Kiribati | 2018 | 3.3 | 6.4 | 9.2 | 5.7 | 6.5 | 4.3 | 4.2 | 13.2 | 8.9 | 7.1 | 3.9 | -4.1; 12.0 |
| Lao | 2017 | 15.1 | 15.1 | 13.6 | 16.0 | 22.4 | 22.8 | 30.9 | 39.2 | 41.2 | 47.7 | 36.8 | 30.7; 42.9 |
| Myanmar | 2015 | 10.8 | 10.8 | 8.6 | 19.3 | 13.0 | 19.0 | 22.4 | 18.7 | 20.6 | 24.2 | 15.6 | 7.1; 24.1 |
| Mongolia | 2018 | 9.0 | 8.5 | 16.0 | 23.0 | 24.0 | 40.6 | 39.7 | 27.8 | 54.6 | 32.8 | 40.3 | 30.3; 50.4 |
| Papua New Guinea | 2016 | 14.5 | 9.5 | 19.2 | 20.0 | 17.2 | 17.5 | 21.8 | 16.5 | 18.2 | 25.3 | 8.4 | 1.0; 15.8 |
| Thailand | 2015 | 57.2 | 50.5 | 52.8 | 48.4 | 65.8 | 54.5 | 45.0 | 55.7 | 49.7 | 54.4 | -0.9 | -16.0; 14.2 |
| Timor Leste | 2016 | 6.4 | 7.4 | 10.8 | 9.0 | 12.4 | 10.9 | 12.1 | 19.9 | 14.5 | 28.8 | 17.8 | 10.6; 25.1 |
| Vietnam | 2013 | 18.9 | 46.6 | 50.8 | 52.6 | 52.2 | 59.3 | 65.6 | 67.3 | 68.2 | 61.0 | 40.0 | 29.7; 50.3 |
| Latin America & Caribbean | | | | | | | | | | | | | |
| Dominican Republic | 2014 | 27.6 | 36.1 | 41.0 | 40.8 | 32.4 | 48.0 | 37.3 | 41.5 | 51.3 | 39.7 | 15.2 | 9.1; 21.2 |
| Ecuador | 2012 | 29.3 | 31.5 | 34.8 | 47.4 | 40.4 | 40.2 | 48.1 | 46.6 | 40.0 | 51.7 | 20.6 | 10.8; 30.3 |
| Guatemala | 2014 | 40.5 | 44.8 | 46.8 | 46.1 | 53.5 | 54.0 | 56.6 | 66.1 | 61.1 | 70.7 | 29.0 | 21.9; 36.1 |
| Guyana | 2014 | 22.7 | 28.9 | 13.0 | 37.1 | 24.4 | 17.4 | 28.1 | 22.6 | 29.2 | 43.6 | 9.4 | -2.6; 21.4 |
| Honduras | 2011 | 37.3 | 51.8 | 54.1 | 53.4 | 52.7 | 52.0 | 68.1 | 64.0 | 60.2 | 64.0 | 25.1 | 17.6; 32.6 |
| Haiti | 2016 | 3.7 | 8.8 | 5.0 | 10.8 | 5.6 | 14.3 | 8.9 | 17.0 | 21.7 | 30.8 | 21.1 | 14.3; 28.0 |
| Mexico | 2015 | 35.8 | 45.4 | 37.6 | 45.9 | 42.4 | 52.3 | 48.2 | 49.1 | 53.1 | 62.8 | 19.3 | 6.4; 32.2 |
| Paraguay | 2016 | 24.0 | 35.6 | 34.8 | 29.6 | 41.3 | 44.0 | 37.6 | 46.3 | 43.0 | 47.3 | 21.9 | 10.9; 32.9 |
| El Salvador | 2014 | 48.3 | 63.2 | 61.8 | 57.7 | 63.6 | 62.6 | 68.1 | 57.8 | 76.4 | 71.8 | 18.7 | 9.3; 28.2 |
| Suriname | 2018 | 7.7 | 7.3 | 10.5 | 21.3 | 18.1 | 9.2 | 16.3 | 7.4 | 11.7 | 14.0 | 5.4 | -2.1; 12.9 |

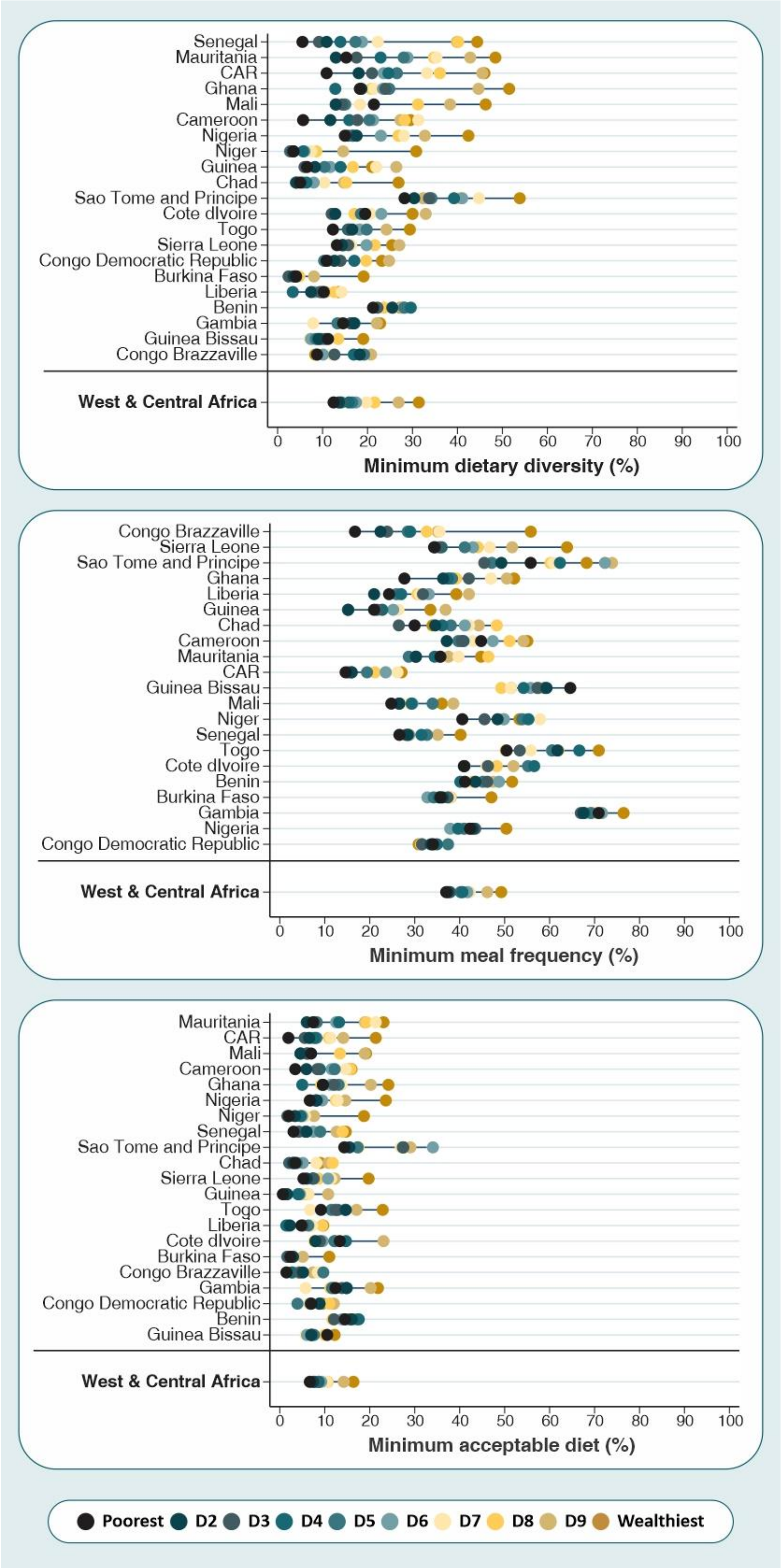


**Supplementary figure 3:** Complementary feeding indicators by wealth deciles for each country from the West & Central Africa region

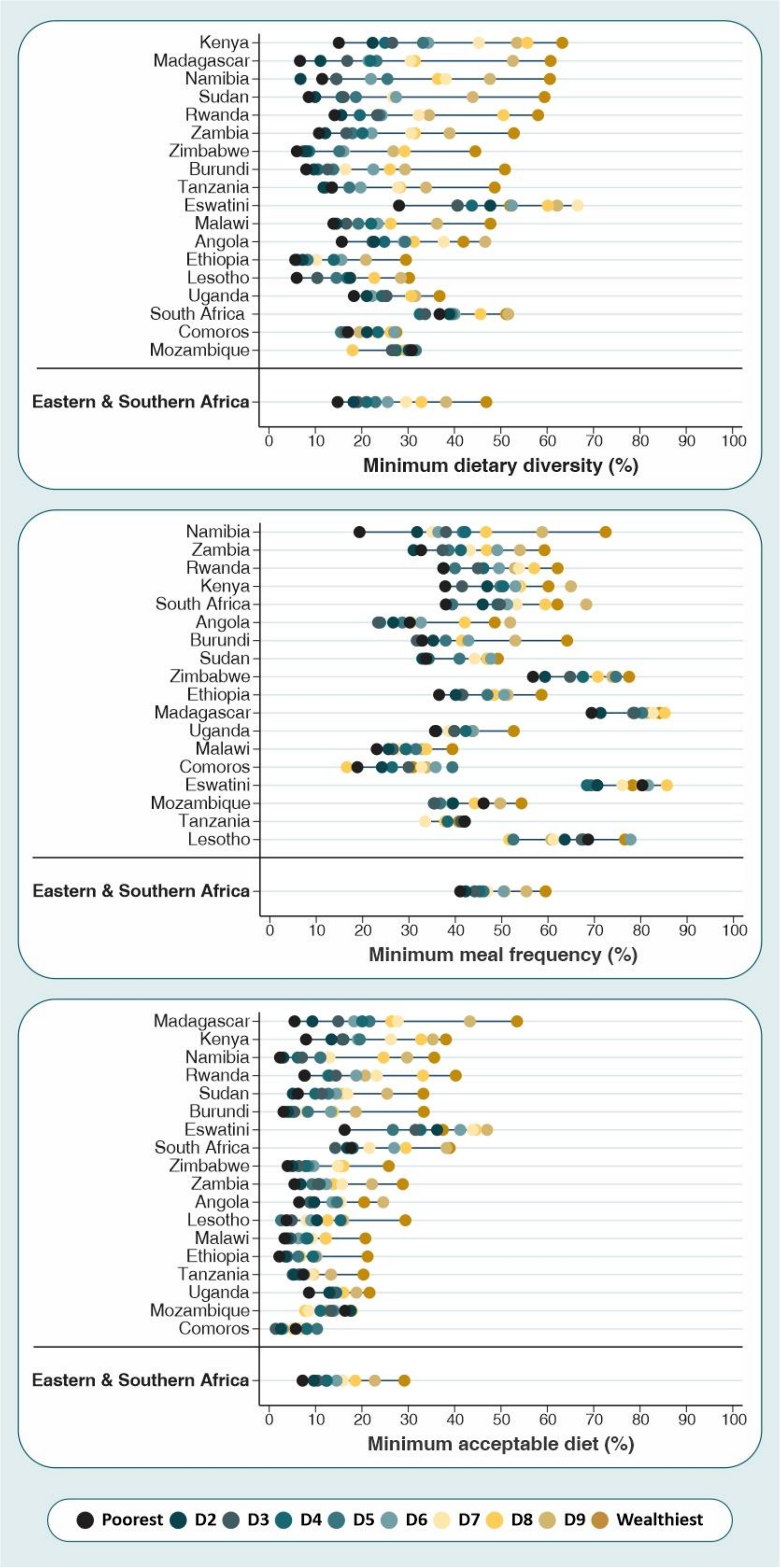


**Supplementary figure 4:** Complementary feeding indicators by wealth deciles for each country from the Eastern & Southern Africa region

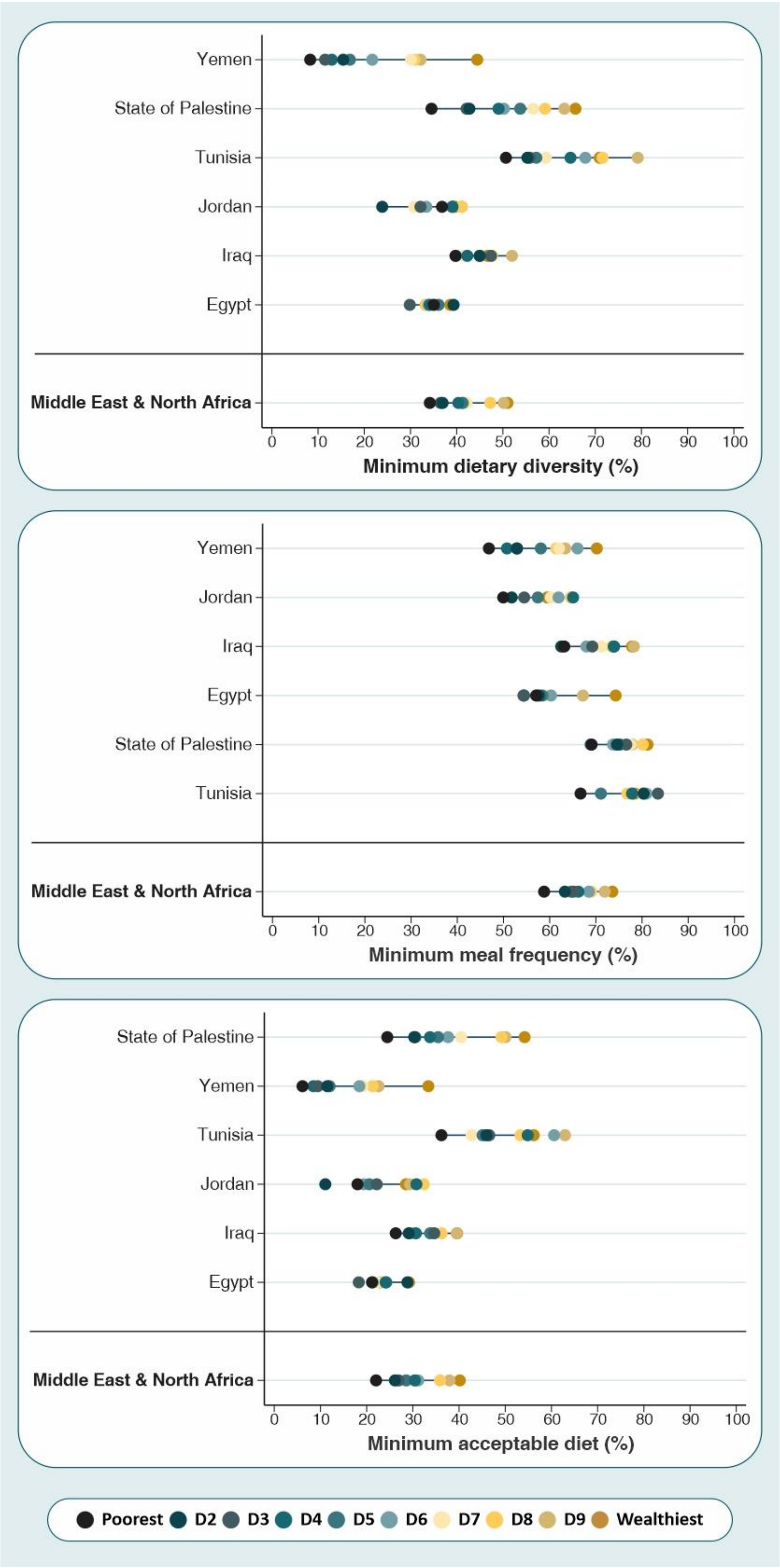


**Supplementary figure 5:** Complementary feeding indicators by wealth deciles for each country from the Middle East & North Africa region

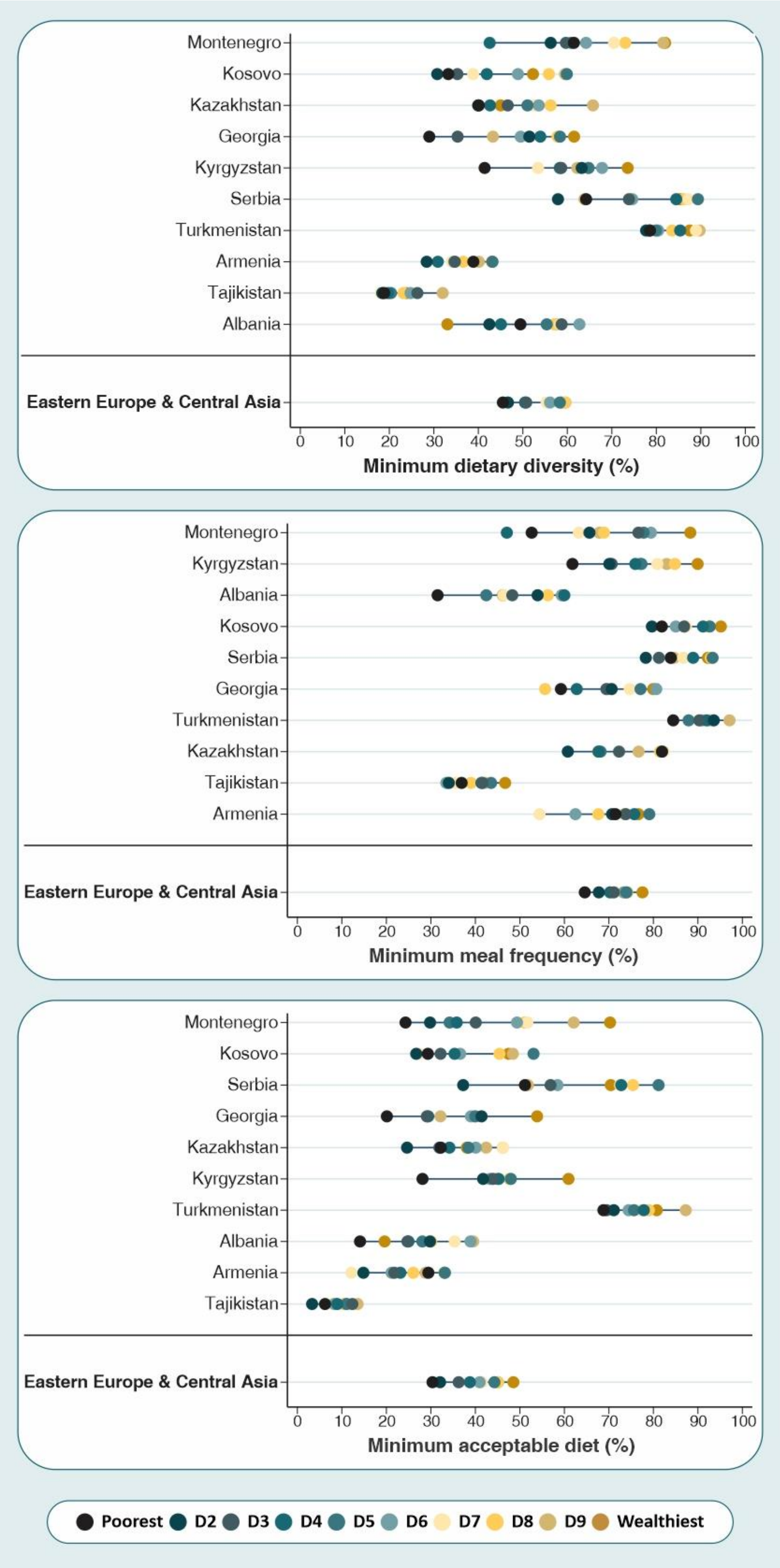


**Supplementary figure 6:** Complementary feeding indicators by wealth deciles for each country from the Eastern Europe & Central Asia region

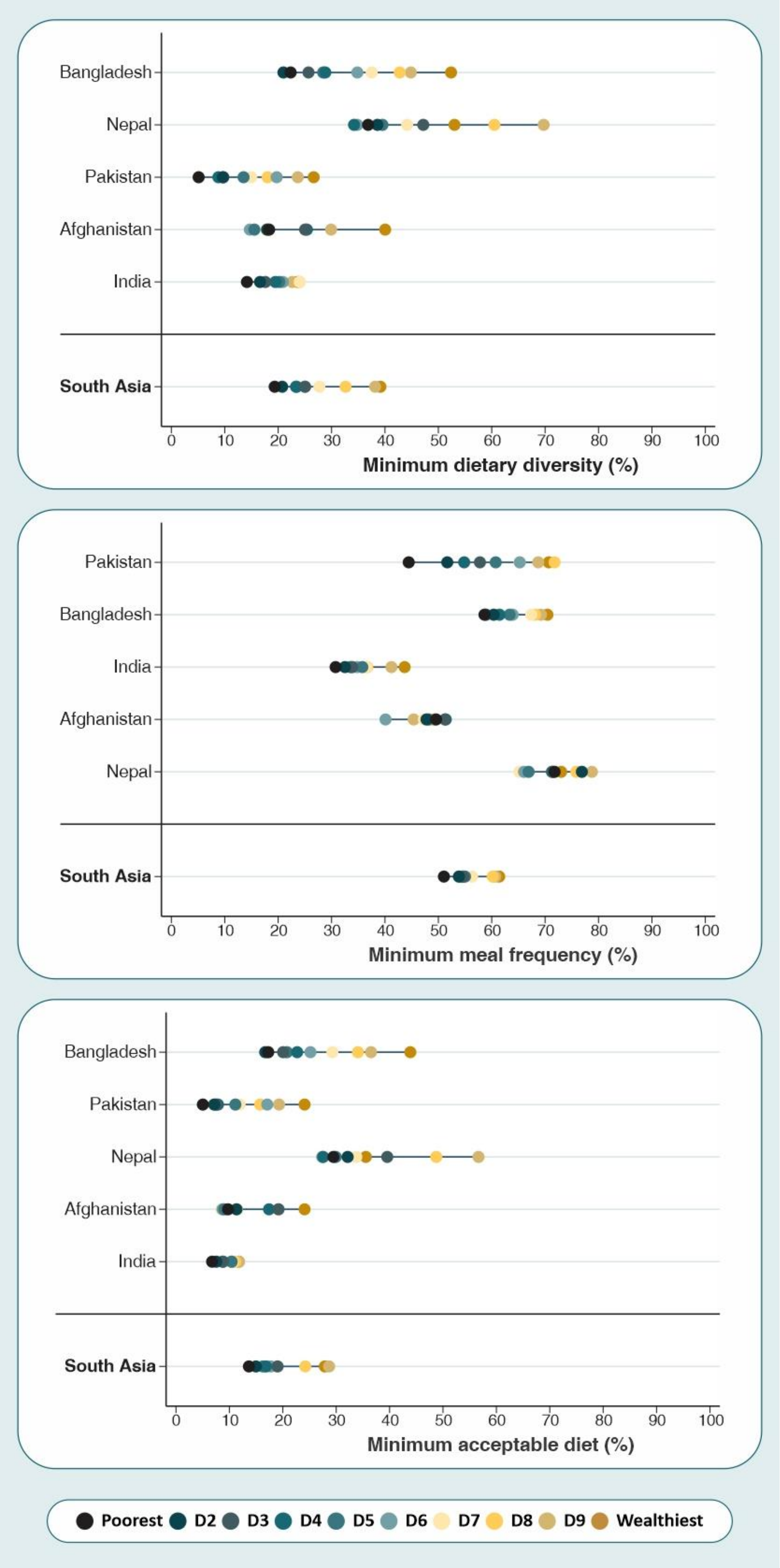


**Supplementary figure 7:** Complementary feeding indicators by wealth deciles for each country from the South Asia region

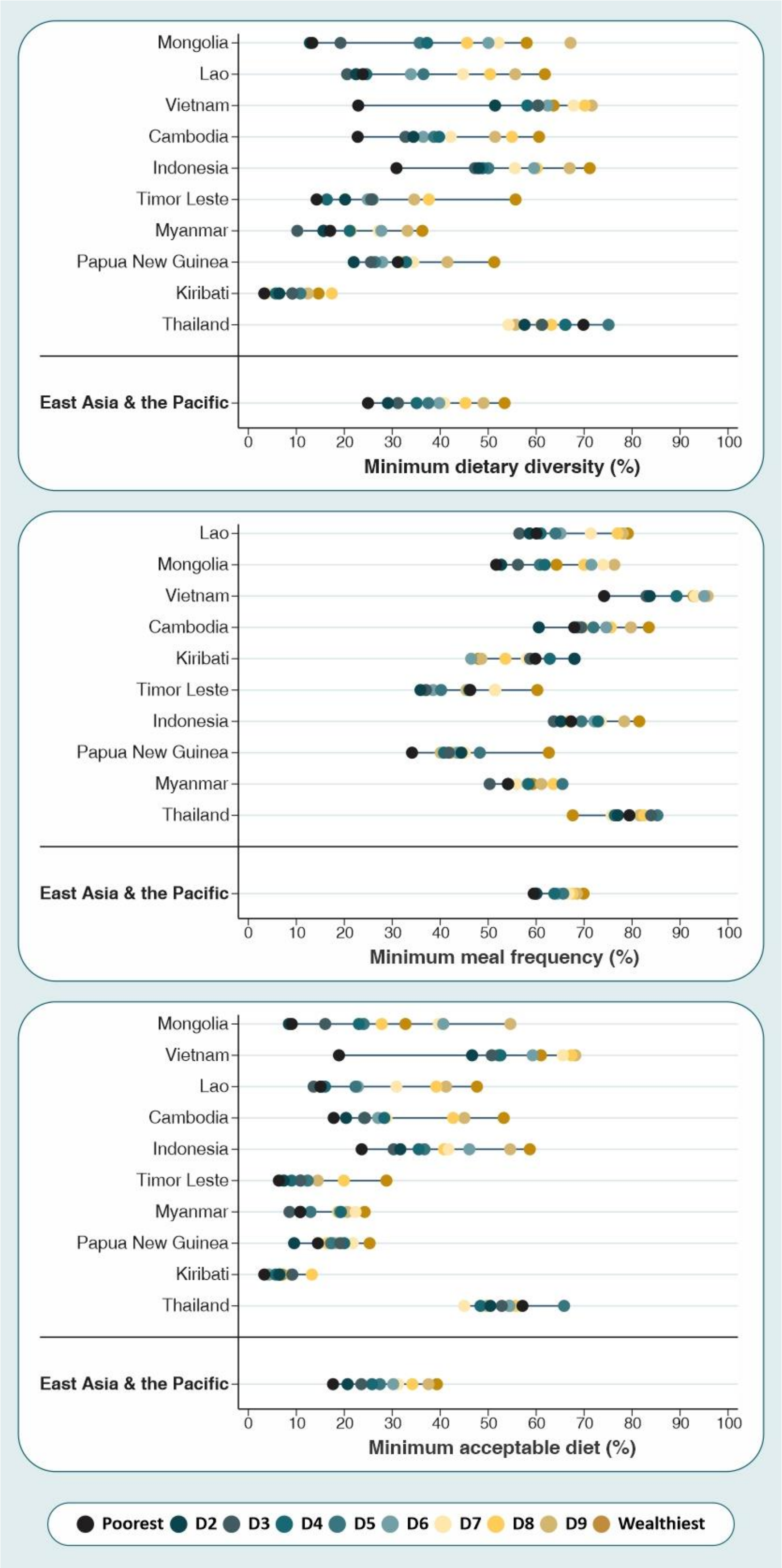


**Supplementary figure 8:** Complementary feeding indicators by wealth deciles for each country from the East Asia & the Pacific region

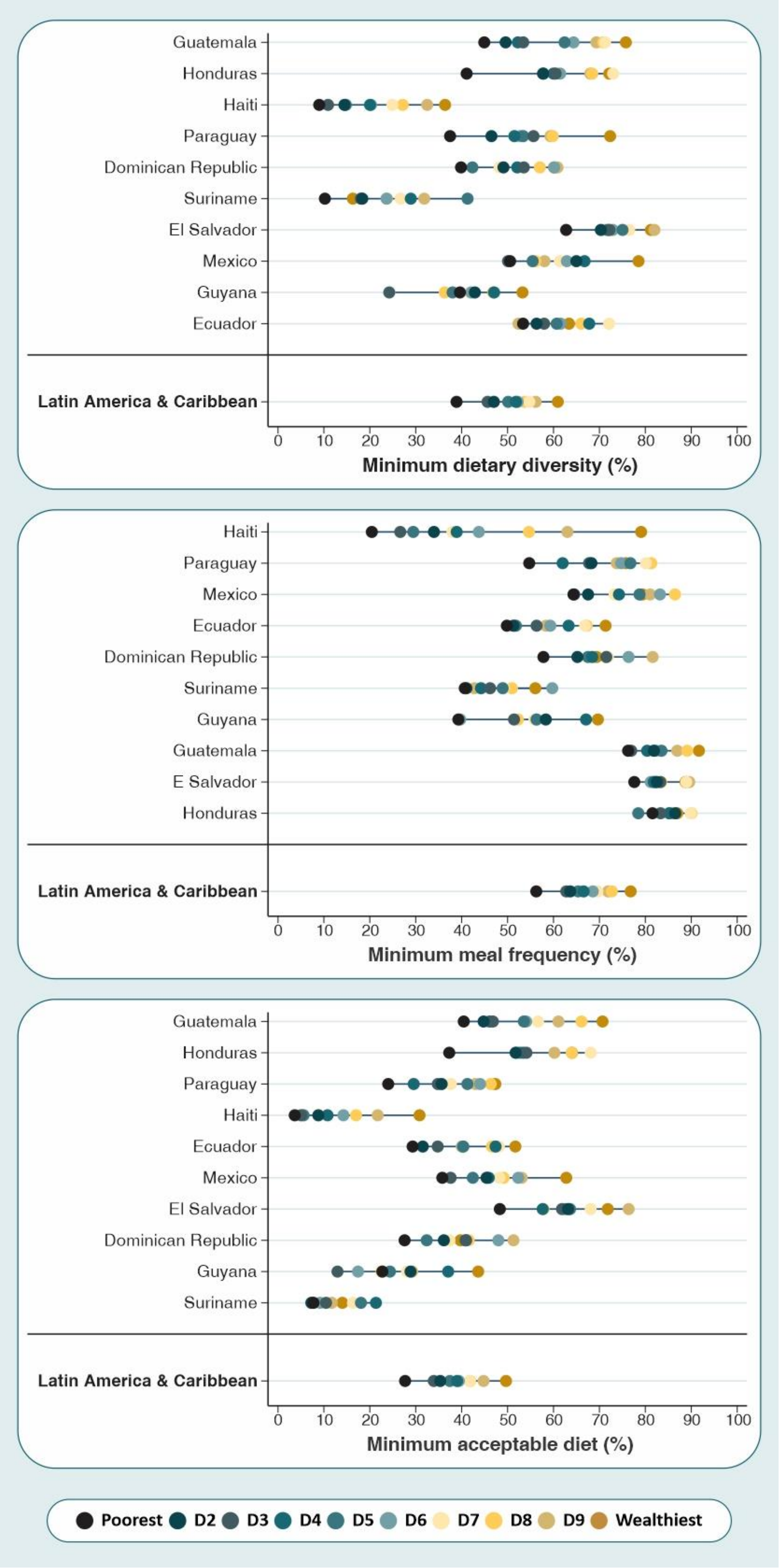


**Supplementary figure 9:** Complementary feeding indicators by wealth deciles for each country from the Latin America & the Caribbean region

**Supplementary table 6**. Mean of the weighted prevalence for each food group by inequality indicators, according to world regions

| Food group | World regions | Poorest decile (%) | Wealthiest decile (%) | SII (pp) | 95% CI |
|---|---|---|---|---|---|
| Breastmilk | West & Central Africa | 34.3 | 26.1 | -8.9 | -10.6; -7.2 |
| | Eastern & Southern Africa | 32.0 | 28.2 | -3.0 | -5.5; -0.4 |
| | Middle East & North Africa | 27.5 | 24.3 | -3.7 | -5.1; -2.4 |
| | Eastern Europe & Central Asia | 25.9 | 23.7 | -1.6 | -4.6; 1.4 |
| | South Asia | 41.0 | 33.1 | -9.0 | -10.5; -7.5 |
| | East Asia & the Pacific | 36.6 | 26.3 | -10.1 | -12.8; -7.4 |
| | Latin America & Caribbean | 31.6 | 16.5 | -17.6 | -19.6; -15.7 |
| | *All regions* | *36.1* | *28.6* | *-8.0* | *-9.4; -6.6* |
| Cereal and grains | West & Central Africa | 33.0 | 35.8 | 3.2 | 2.5; 4.0 |
| | Eastern & Southern Africa | 32.2 | 31.7 | -1.1 | -2.1; -0.1 |
| | Middle East & North Africa | 43.8 | 47.0 | 4.0 | 1.7; 6.4 |
| | Eastern Europe & Central Asia | 38.6 | 39.5 | 1.0 | -2.2; 4.2 |
| | South Asia | 28.8 | 26.9 | -1.2 | -3.7; 1.2 |
| | East Asia & the Pacific | 37.9 | 39.0 | 2.1 | -0.1; 4.4 |
| | Latin America & Caribbean | 48.3 | 52.0 | 0.2 | -8.5; 8.8 |
| | *All regions* | *33.2* | *33.4* | *0.5* | *-0.1; 1.0* |
| Legumes and nuts | West & Central Africa | 9.7 | 9.1 | -1.0 | -2.1; 0.2 |
| | Eastern & Southern Africa | 10.0 | 12.9 | 3.0 | 2.1; 3.8 |
| | Middle East & North Africa | 13.2 | 11.7 | -2.0 | -5.2; 1.2 |
| | Eastern Europe & Central Asia | 4.2 | 6.4 | 2.1 | 0.7; 3.4 |
| | South Asia | 4.9 | 5.6 | 1.0 | 0.4; 1.6 |
| | East Asia & the Pacific | 6.1 | 11.3 | 5.4 | 3.9; 6.9 |
| | Latin America & Caribbean | 30.0 | 22.8 | -7.0 | -11.3; -2.6 |
| | *All regions* | *8.6* | *9.4* | *0.9* | *0.1; 1.7* |
| Dairy products | West & Central Africa | 9.6 | 26.9 | 18.0 | 10.4; 25.6 |
| | Eastern & Southern Africa | 14.5 | 24.2 | 11.3 | 4.6; 18.0 |
| | Middle East & North Africa | 41.7 | 47.6 | 7.9 | 4.2; 11.6 |
| | Eastern Europe & Central Asia | 34.2 | 38.2 | 2.9 | -0.5; 6.3 |
| | South Asia | 17.0 | 27.6 | 11.2 | 9.2; 13.3 |
| | East Asia & the Pacific | 13.3 | 37.2 | 22.7 | 18.6; 26.7 |
| | Latin America & Caribbean | 32.8 | 51.9 | 15.3 | 6.5; 24.2 |
| | *All regions* | *17.1* | *30.3* | *13.7* | *11.6; 15.7* |
| Flesh foods | West & Central Africa | 11.5 | 24.3 | 13.7 | 13.0; 14.5 |
| | Eastern & Southern Africa | 10.4 | 16.0 | 5.1 | 3.9; 6.4 |
| | Middle East & North Africa | 18.5 | 26.5 | 9.2 | 7.8; 10.7 |
| | Eastern Europe & Central Asia | 24.5 | 26.1 | 2.3 | -1.1; 5.7 |
| | South Asia | 4.3 | 5.2 | 1.1 | 0.3; 1.9 |
| | East Asia & the Pacific | 21.3 | 32.6 | 10.6 | 8.5; 12.7 |
| | Latin America & Caribbean | 23.0 | 41.7 | 15.2 | 5.9; 24.5 |
| | *All regions* | *10.7* | *17.2* | *6.6* | *5.9; 7.3* |

| **Food group** | **World regions** | **Poorest decile (%)** | **Wealthiest decile (%)** | **SII** | **95% CI** |
|---|---|---|---|---|---|
| Eggs | West & Central Africa | 2.5 | 13.8 | 12.2 | 10.2; 14.2 |
| | Eastern & Southern Africa | 3.9 | 10.6 | 6.5 | 4.7; 8.3 |
| | Middle East & North Africa | 16.9 | 17.5 | 1.0 | -0.8; 2.9 |
| | Eastern Europe & Central Asia | 14.0 | 14.0 | -0.3 | -2.3; 1.8 |
| | South Asia | 5.4 | 8.3 | 4.0 | 2.3; 5.8 |
| | East Asia & the Pacific | 14.3 | 22.5 | 5.7 | 2.7; 8.7 |
| | Latin America & Caribbean | 19.6 | 27.4 | 1.7 | -4.6; 8.0 |
| | *All regions* | *6.9* | *12.8* | *5.8* | *5.1; 6.6* |
| Vitamin A rich fruits and vegetables | West & Central Africa | 20.3 | 20.4 | -0.8 | -2.4; 0.8 |
| | Eastern & Southern Africa | 19.8 | 20.5 | -0.1 | -1.0; 0.9 |
| | Middle East & North Africa | 16.8 | 22.5 | 4.9 | 2.9; 6.9 |
| | Eastern Europe & Central Asia | 18.7 | 23.1 | 4.9 | 1.8; 7.9 |
| | South Asia | 14.6 | 13.5 | -1.2 | -1.7; -0.6 |
| | East Asia & the Pacific | 26.4 | 35.7 | 9.5 | 7.5; 11.5 |
| | Latin America & Caribbean | 23.2 | 34.9 | 10.9 | 4.5; 17.4 |
| | *All regions* | *18.5* | *20.2* | *1.2* | *0.5; 1.9* |
| Other fruits and vegetables | West & Central Africa | 5.9 | 11.0 | 4.9 | 3.6; 6.2 |
| | Eastern & Southern Africa | 5.9 | 13.5 | 7.4 | 6.2; 8.6 |
| | Middle East & North Africa | 19.5 | 28.5 | 8.7 | 6.9; 10.5 |
| | Eastern Europe & Central Asia | 24.1 | 31.0 | 7.0 | 3.1; 10.8 |
| | South Asia | 7.1 | 11.3 | 5.0 | 3.1; 6.8 |
| | East Asia & the Pacific | 8.0 | 18.0 | 9.0 | 6.9; 11.1 |
| | Latin America & Caribbean | 28.3 | 43.3 | 13.1 | 3.9; 22.3 |
| | *All regions* | *8.7* | *15.1* | *6.4* | *5.6; 7.2* |
